# Mindfulness-based intervention in preterm young adolescents: benefits on neurobehavioural functioning and its association with white-matter microstructural changes

**DOI:** 10.1101/2021.10.20.21265246

**Authors:** Vanessa Siffredi, Maria Chiara Liverani, Dimitri Van De Ville, Lorena G. A. Freitas, Cristina Borradori Tolsa, Petra Susan Hüppi, Russia Hà-Vinh Leuchter

## Abstract

Very preterm (VPT) young adolescents are at high risk of executive, behavioural and socio-emotional difficulties. Previous research has shown significant evidence of the benefits of mindfulness-based intervention (MBI) on these abilities. This study aims to assess the association between the effects of MBI on neurobehavioral functioning and changes in white-matter microstructure in VPT young adolescents who completed an 8-week MBI program. Neurobehavioural assessments and multi-shell diffusion MRI were performed before and after MBI in 32 VPT young adolescents. Combined diffusion tensor imaging (DTI) and neurite orientation dispersion and density imaging (NODDI) measures were extracted on well-defined white matter tracts (TractSeg). A multivariate data-driven approach (partial least squares correlation) was used to explore associations between MBI-related changes on neurobehavioural measures and microstructural changes. Our finding showed an enhancement of global executive functioning after MBI that was associated with a general pattern of significant increase in fractional anisotropy (FA) and decrease in axonal dispersion (ODI) in white-matter tracts involved in executive processes. Young VPT adolescents with lower gestational age at birth showed the greatest gain in white-matter microstructural changes after MBI.

**Highlights:**

- Very preterm adolescents (VPT) completed an 8-week mindfulness-based intervention (MBI)
- Improvement in overall executive functioning was observed after MBI
- Executive gain was associate with white-matter microstructural changes
- The increase in microstructural properties was in tracts involved in executive processes
- VPT with lower gestational age show bigger gains in microstructural changes

**CRediT roles:** **Vanessa Siffredi:** Conceptualization; Data curation; Formal analysis; Investigation; Methodology; Project administration; Software; Visualization; Writing - original draft; Writing - review & editing - **Maria Chiara Liverani:** Conceptualization; Data curation; Investigation; Methodology; Project administration; Writing - review & editing. **- Dimitri Van De Ville**: Methodology; Resources; Software; Supervision; Writing - review & editing. **- Lorena Freitas:** Data curation; Investigation; Writing - review & editing. - **Cristina Borradori Tolsa:** Conceptualization; Funding acquisition; Investigation; Project administration; Resources; Supervision; Validation; Writing - review & editing. - **Petra Susan Hüppi:** Conceptualization; Funding acquisition; Methodology; Project administration; Resources; Supervision; Validation; Writing - review & editing. -**Russia Hà-Vinh Leuchter:** Conceptualization; Funding acquisition; Investigation; Methodology; Project administration; Resources; Supervision; Validation; Writing - review & editing.

## Introduction

Children and adolescents born very preterm (VPT; <32 completed weeks of gestation) are at increased risk for executive, behavioural and socio-emotional impairments that persist into adolescence and adulthood. Executive functioning (EF) is essential for goal-directed and adaptive problem-solving and behaviour. It has been conceptualised in four distinct subdomains: (i) attentional control, (ii) information processing, (iii) cognitive flexibility, and (iv) goal setting (Anderson, 2002). Behavioural and socio-emotional competences refer to a set of skills related to how individuals identify, express, understand, use and regulate their behaviours as well as their emotions and those of others (Mikolajczak, Quoidbach, Kotsou, & Nelis, 2009). Importantly, these competences are crucial in daily life activities and are closely linked to academic abilities and social behaviours (Best, Miller, & Naglieri, 2011; Carter, Briggs-Gowan, & Davis, 2004; Izard, 2011; Vaughan & Giovanello, 2010). Numerous studies show that white matter alterations are highly common after preterm birth and persist until adolescence and adulthood (Volpe, 2003). Importantly, these white-matter changes have been associated with executive, behavioural and socioemotional deficits in this population (Allin et al., 2011; K. M. Jones, Champion, & Woodward, 2013; Loe, Lee, & Feldman, 2013; Montagna & Nosarti, 2016; Nosarti, Allin, Frangou, Rifkin, & Murray, 2005; Perlman, 1998; Soria-Pastor et al., 2008; Thompson et al., 2014; Woodward, Clark, Pritchard, Anderson, & Inder, 2011).

Mindfulness-based intervention (MBI) - commonly defined as the ongoing monitoring of present-moment experience while attending to it in an open and accepting way and without judgment (Kabat-Zinn, 2003) - has been associated with enhanced executive, behavioural and socioemotional functioning in typically developing children and adolescents (Barnes, Bauza, & Treiber, 2003; Beauchemin, Hutchins, & Patterson, 2008; Biegel, Brown, Shapiro, & Schubert, 2009; Black, Milam, & Sussman, 2009; Broderick & Metz, 2009; Felver, Tipsord, Morris, Racer, & Dishion, 2017; Flook et al., 2010; Geronimi, Arellano, & Woodruff-Borden, 2019; Lee, Semple, Rosa, & Miller, 2008; Napoli, Krech, & Holley, 2004; Schonert-Reichl & Lawlor, 2010; Schonert-Reichl et al., 2015; Semple, Lee, Rosa, & Miller, 2009). In line with behavioural improvements, studies show that MBI may induce structural neuroplastic changes (Marchand, 2014). In this context, diffusion-weighted magnetic resonance imaging (DW-MRI) and the diffusion tensor imaging (DTI) model has been shown to be sensitive to subtle white matter microstructural changes. Fractional anisotropy (FA) has been shown to reflect functionally relevant microstructural properties of white matter, including axonal architecture, extent of myelination and density of axonal fibres comprising axonal bundles (C. Beaulieu, 2002; Boorman, O’Shea, Sebastian, Rushworth, & Johansen-Berg, 2007). Increased FA measures have been associated with MBI in adult populations in different brain regions and tracts, including frontal regions (Kang et al., 2013; Luders, Clark, Narr, & Toga, 2011), callosal regions (Laneri et al., 2016; Luders et al., 2012), anterior cingulate regions (Tang, Lu, Fan, Yang, & Posner, 2012), insular (Sharp et al., 2018), ucinate fasciculus (Luders et al., 2011) and superior longitudinal fasciculus (Hölzel et al., 2016; Luders et al., 2011). In adolescents, a recent study also showed increased FA in the superior longitudinal fasciculus after a 2-weeks mindfulness video game (Patsenko et al., 2019). To our knowledge, the DTI model is the only methods employed so far to explore potential structural neuroplasticity induced by MBI.

We recently reported a randomised controlled trial showing significant benefits of MBI on executive, behavioural and socio-emotional functioning in preterm young adolescents (Siffredi, Liverani, Hüppi, et al., 2021). The current study aims to assess the association between the benefits of MBI on neurobehavioral functioning and changes in microstructural brain properties. To this end, we first compared preterm and full-term young adolescents on a range of executive, behavioural and socio-emotional measures. Secondly, we assessed the benefit of an 8-weeks MBI on the neurobehavioural measures in which preterm young adolescents showed significant difficulties (i.e., significant reduction in neurobehavioural scores compared to the full-term group). Finally, we explored the association between changes on neurobehavioural measures after MBI and changes on white-matter microstructural measures. As described above, previous findings have revealed changes in quantitative DTI metrics after MBI, specifically an increase in FA. One potential confound of the DTI analyses relates to the inherent non-specificity of the measurement, where observed changes in FA may be due to changes to any combination of axonal dispersion, axonal densities or fibre crossing (Christian Beaulieu, 2002; D. K. Jones, Knösche, & Turner, 2013). In an attempt to provide more specific information about white-matter microstructural changes associated with MBI, a biophysical model of diffusion, NODDI (neurite orientation dispersion and density imaging), was used. The NODDI model allows to distinguish two key variables contributing to changes in FA, including neurite density and fibre orientation dispersion (Zhang, Schneider, Wheeler-Kingshott, & Alexander, 2012). Combining the high sensitivity of the DTI model with the high specificity of the NODDI model might allow for a better understanding of white-matter microstructural changes associated with changes on neurobehavioural measures after MBI.

## Methods

### Participants

This study uses data collected as part of the ‘Mindful preterm teens’ study (Siffredi, Liverani, Magnus-Smith, et al., 2021). One hundred and sixty-five VPT young adolescents, i.e., born before 32 gestational weeks, were invited to participate in the study. They were aged 10 to 14 years born between 01.01.2003 and 31.12.2008 at the Geneva University Hospital, Switzerland, and followed at the Division of Child Development and Growth at the Geneva University Hospital. VPT young adolescents were excluded if they had an intelligence quotient below 70, sensory or physical disabilities (cerebral palsy, blindness, hearing loss), or an insufficient understanding of French. Moreover, some families declined to participate due to lack of time, lack of interest, geographical constraints or unreachability. A total of 63 participants were enrolled in the ‘Mindful preterm teens’ study and 52 of them completed the MBI and neurobehavioural assessment before and after MBI. Of the 52 young adolescents, 39 completed MRI scans before and after intervention, and 32 were included in the current diffusion MRI analyses (diffusion sequences not completed, n=5; high level of motion artefacts, n=2), Figure 1.

**Figure 1.**
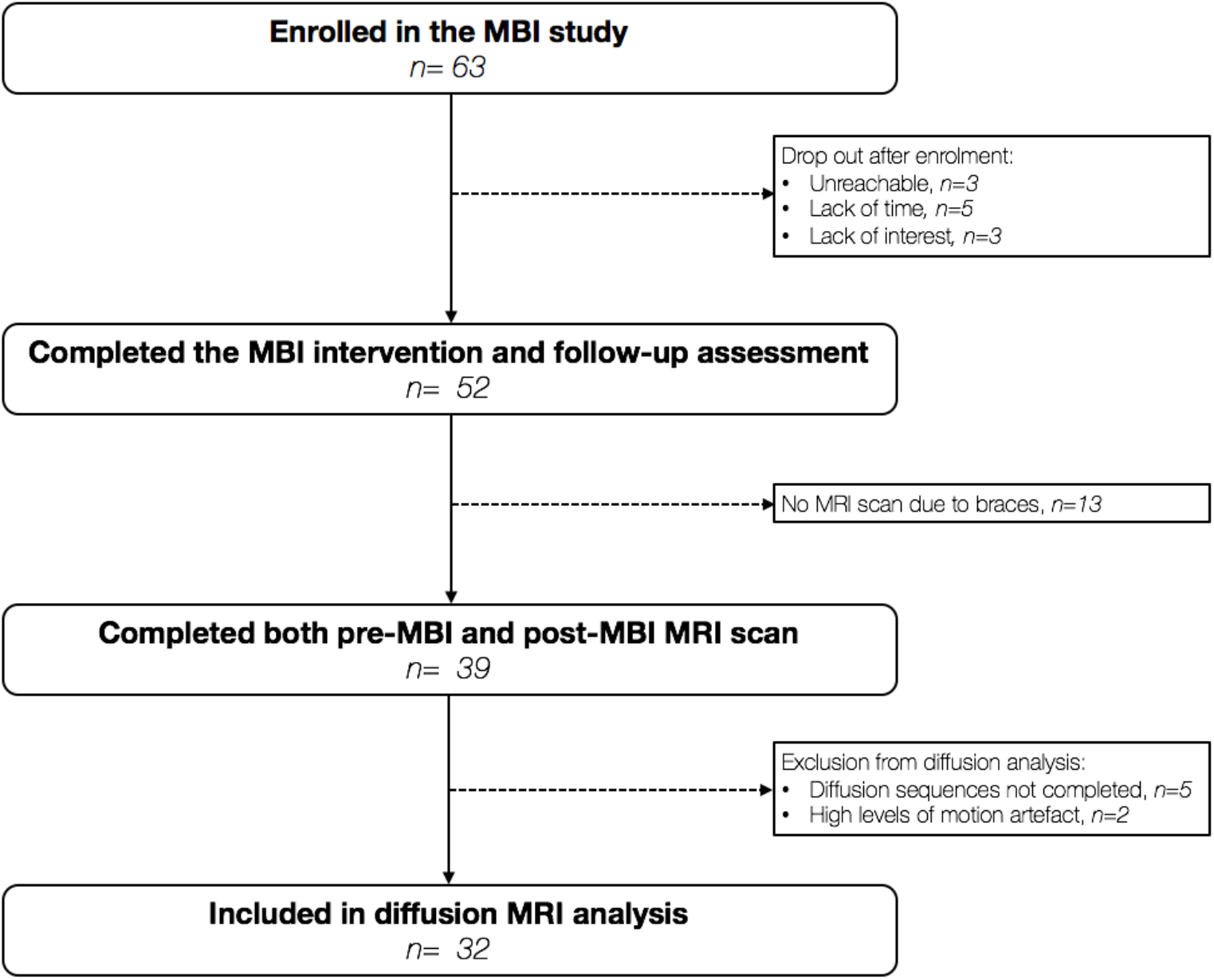
Participant flow chart. This figure shows the number of young adolescents who were enrolled into the MBI study, and of these, the number who had usable scans at both pre- and post-MBI time points and were thus included in the current analysis (Siffredi, Liverani, Magnus-Smith, et al., 2021). MBI, Mindfulness-based intervention; MRI, magnetic resonance imaging.

Moreover, 22 term-born young adolescents aged between 10 and 14 years old were recruited through the community and completed a neurobehavioural assessment similar to the VPT group prior to the MBI. This study was approved by the Swiss Ethics Committees on research involving humans, ID: 2015-00175. Written informed consent was obtained from the principal caregiver and from the participant.

### Mindfulness-based intervention

The proposed MBI was designed by the authors, adapting well-known MBI programs, including Mindfulness-Based Stress Reduction (MBSR; (Kabat-Zinn, 2003)) and Mindfulness-Based Cognitive Therapy (MBCT; (Segal, Williams, & Teasdale, 2001)), to adolescents’ needs and language. The length of sessions and practices were also adapted to this population. The program consisted of 8 weekly sessions in groups of up to 8 participants, lasting 1h30. Two instructors were present for each group throughout the intervention (for further details on the MBI, see (Siffredi, Liverani, Magnus-Smith, et al., 2021)).

### Neurobehavioral measures

Participants’ executive, behavioural and socio-emotional functioning were assessed using parent-report and self-report questionnaires, neuropsychological testing and computerised neurocognitive tasks, see Supplementary Table S1.

#### (i) Executive competences measures

Behaviours related to executive competences of young adolescents were assessed using the Behaviour Rating Inventory of Executive Function-parent version (BRIEF) (Gioia, Isquith, Guy, & Kenworthy, 2000). The BRIEF comprises 86 items over two standardised subscales, the Behavioural Regulation Index (BRI), which evaluates inhibition, flexibility and emotional control, and the Metacognition Index (MI), which evaluates initiation, working memory, planning/organization, self-regulation as well as a global score called the Global Executive Composite (GEC). Neurocognitive computerised tasks comprised: (i) the Flanker Visual Filtering Task, in which reaction time of the congruent condition was used to assess speed of processing (which belongs to the information processing subdomain), and the inhibition score (reaction time in incongruent conditions – reaction time in congruent conditions) was used as a measure of the subdomain of attentional control (Anderson, 2002; Christ, Kester, Bodner, & Miles, 2011); (ii) the child-adapted version of the Reality Filtering task, in which the temporal context confusion index (TCC) was used as a reality filtering measure, which involves integration of different executive processes (Liverani et al., 2020; Liverani et al., 2017). Neuropsychological testing included the Letter-Number Sequencing subtest from the Wechsler Intelligence Scale for Children, 4^th^ Edition (WISC-IV) assessing working memory, which belongs to the cognitive flexibility subdomain (Anderson, 2002). Given the strong association between executive functions and mathematical abilities in children and adolescents,(Holmes & Adams, 2006; Visu-Petra, Cheie, Benga, & Miclea, 2011) we also used the total score of the Tempo Test Rekenen to assess timed mathematical achievement (De Vos, 1994).

#### (ii) Behavioural and socio-emotional competences measures

The total score of the Strength and Difficulties Questionnaire – parent version (SDQ) was used to assess behaviour in daily life (Goodman, 1997, 2001). Participants completed three self-reported questionnaires: the KIDSCREEN-27 items questionnaire was used to assess the quality of life of the participants (Robitail et al., 2007); the total score of the Social Goal Scale was used to assess social responsiveness and social relationships (Wentzel, 1994); and the total score of the Self-Compassion Scale – Short form was used to assess the main components of self-compassion (Raes, Pommier, Neff, & Van Gucht, 2011). Neuropsychological testing included the Affect Recognition subtest (A Developmental Neuropsychological Assessment, 2nd Edition - NEPSY-II), giving a total score assessing facial emotional recognition and the Theory of Mind subtest (NEPSY-II), giving a total score measuring the ability to understand mental functions, such as belief, intention or deception (Korkman, Kirk, & Kemp, 2007).

### Magnetic resonance imaging

MRI data were acquired at Campus Biotech in Geneva, Switzerland, using a Siemens 3T Magnetom Prisma scanner. All participants completed a simulated “mock” MRI session prior to their first MRI scan. This preparation process was conducted by trained research staff and allowed participants to familiarise themselves with the scanner and the scanning process, eventually raising any concerns they might have had prior to the MRI scan. Furthermore, this process is known to facilitate the acquisition of good quality MRI images in children and adolescents (de Bie et al., 2010; Tamnes, Roalf, Goddings, & Lebel, 2018).

A multi-shell diffusion-weighted (DW) echo planar imaging (EPI) protocol was used and included four shells. The first sequence, referred to as ‘*b200*’, included 10 gradient directions with b-values of 200 s/mm2; the second one referred to as ‘*b1700*’, included 30 gradient directions with b-values of 1700 s/mm2; the third one referred to as ‘*b4200a*’, included 26 gradient directions with b-values of 4200 s/mm2; and the fourth one referred to as ‘*b4200b*’, included 24 gradient directions with b-values of 4200 s/mm2. Each of four sequences included the acquisition of 4 images with b-value = 0 s/mm2 images, and the parameter set for all sequences were: TR = 7000 ms, TE = 87 ms, FOV = 234 × 243 mm, slice thickness = 1.3 mm, voxel size = 1.3 × 1.3 × 1.3 mm.

### Diffusion image preprocessing

Visual inspection of raw data for brain coverage, spike artefacts, severe head motion, and other severe image artefacts was completed and participants were excluded if necessary. The four diffusion shells (*b200, b1700, b4200a, b4200b*) were preprocessed independently using MRtrix3 (Tournier et al., 2019) and using the following pipeline: a) denoising (Cordero-Grande, Christiaens, Hutter, Price, & Hajnal, 2019; Veraart, Fieremans, & Novikov, 2016; Veraart, Novikov, et al., 2016);, b) Gibbs ringing removal (Kellner, Dhital, Kiselev, & Reisert, 2016), c) correction for movement and eddy current-induced geometric distortions using the eddy tool implemented in FSL (Jenkinson, Beckmann, Behrens, Woolrich, & Smith, 2012). The first b = 0 s/mm2 images of the *b1700, b4200a, b4200b* sequences were linearly registered to the first b = 0 s/mm2 image of the *b200* sequence using FreeSurfer to bring them into *b200* space before merging them together. The brain extraction tool (BET) from FSL (Smith, 2002) was then applied to the combined *b200, b1700, b4200a, b4200b* image to remove non-brain tissue and subsequently intensity normalisation was applied. Following Pines and colleague’s (2020) recommendations (Pines et al., 2020), the resulting multi-shell diffusion weighted image was then used for both DTI and NODDI models fitting and tractography.

### Diffusion models fitting

The DTI model was applied to the resulting multi-shell diffusion weighted image and whole-brain maps of FA was calculated for each participant. FA (between 0 and 1) is a measure of the directionality of diffusion that characterise the variance of the three eigenvalues pairs that represent the direction and magnitude of diffusivity along the three orthogonal axes (v1, λ1; v2, λ2; v3, λ3) (Tamnes et al., 2018). In addition, intra-cellular volume fraction and orientation dispersion indices were estimated from the resulting multi-shell diffusion weighted image using the NODDI model (Zhang et al., 2012). The NODDI Matlab Toolbox http://www.nitrc.org/projects/noddi_toolbox was used to extract maps of intra-cellular volume fraction (ICVF) and fibre orientation dispersion (ODI) across the brain for each participant.

### Tractography and tractometry measures

Whole-brain fibre orientation distributions (FOD) were estimated using the multi-shell multi-tissue constrained spherical deconvolution (MSMT-CSD) method (Jeurissen, Tournier, Dhollander, Connelly, & Sijbers, 2014), resulting in a condensed representation of diffusion along three principal fibre directions per voxel according to tissue type (grey, white, cortico-spinal fluid). Tractography-based Segmentation (TractSeg) use a supervised-learning approach with a aconvolutional neural network-based that directly segments tracts in the field of fibre orientation distribution function (fODF) peaks without using parcellation (Wasserthal, Neher, & Maier-Hein, 2018). TractSeg has achieved state-of-the-art performance and allows for an accurate reconstruction of fibre tracts in subject space, thus avoiding the problem of inaccurate coregistration of tracts with varying size and shape. Whole-brain fibre orientation distribution function (fODF) peaks map were input into a two stage fully convolutional neural network trained using segmented priors of 72 anatomically well-defined white matter tracts from the Human Connectome Project. Using the tractometry function, along-tract mean FA, ICVF and ODI were calculated for the 50 most consistent white-matter tracts in each subject’s native space (Chandio, Harezlak, & Garyfallidis, 2019), see Figure 2.

**Figure 2.**
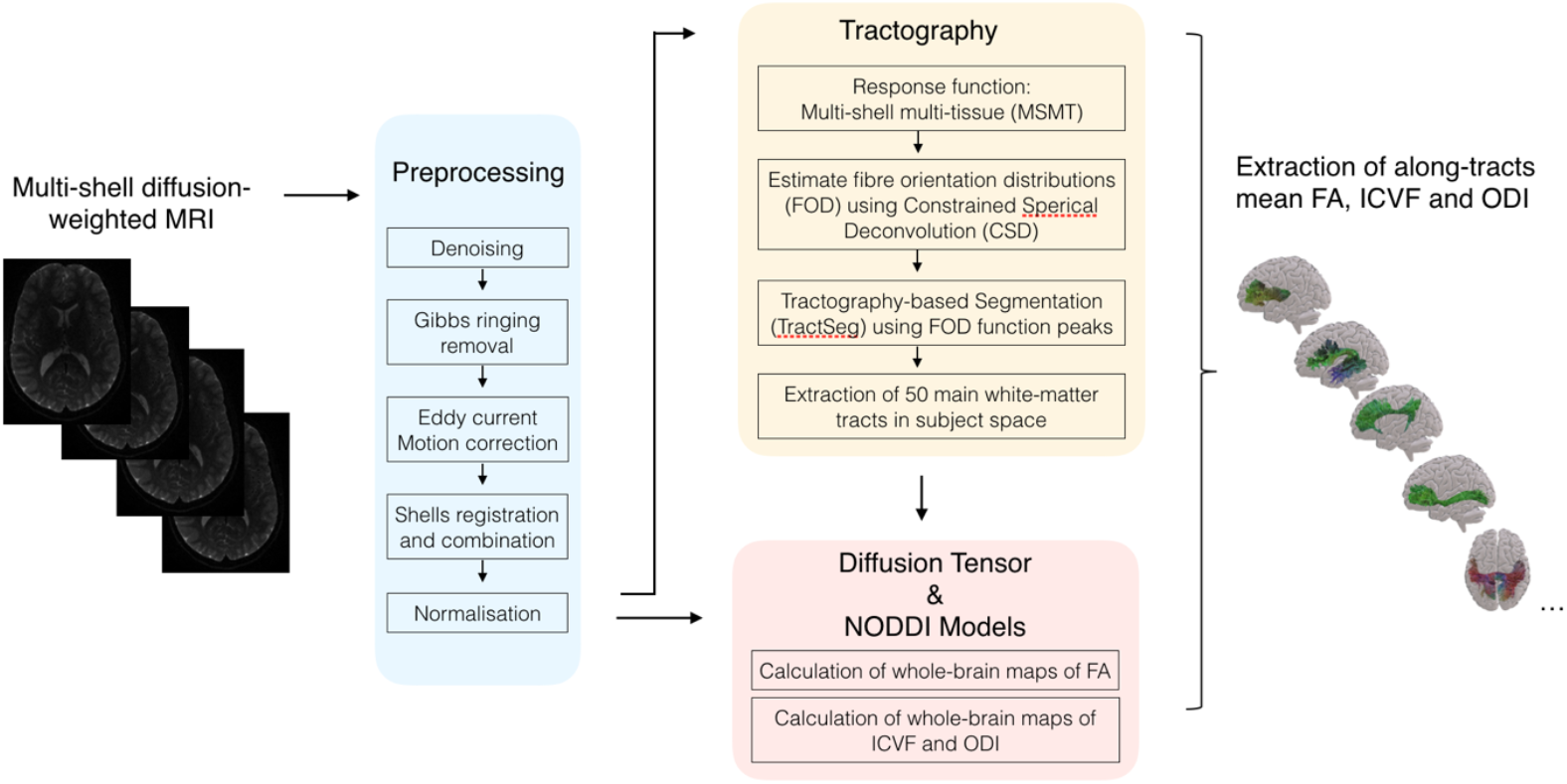
Flowchart summarizing the computation and analysis of microstructural properties of white-matter tracts. Briefly, diffusion-weighted images were preprocessed. Whole-brain multi-shell multi-tissue constrained spherical deconvolution (MSMT-CSD) was extracted and fibre orientation distribution (FOD) function peaks map were input into Tractography-based Segmentation (TractSeg) to extract the main white-matter tracts in subject space. Mean fractional anisotropy (FA), intra-cellular volume fraction (ICVF) and orientation dispersion index (ODI) were then extracted along each track by applying the diffusion tensor and NODDI models.

### Statistical analyses

Statistical analyses were performed using R version 4.0.3 (R. C. Team, 2019) and RStudio version 1.3.1093 (R. Team, 2020) and employing the following methods:

#### Neurobehavioural functioning: comparison of the preterm and full-term groups

Group comparisons between the preterm and the full-term groups were conducted for all neurobehavioural measures using Wilcoxon signed rank test, given that assumptions for parametric testing were violated. P values were corrected for multiple comparisons with the false discovery rate (FDR, q values <0.05 (Benjamini & Hochberg, 1995)). Effect size were assessed using Wilcoxon effect size (r).

#### Impact of MBI on neurobehavioral functioning affected by preterm-birth

Paired-sample t-test were used to evaluate the impact of MBI on the neurobehavioural measures showing a significant deficit in the preterm group as compared to the full-term group. P values were corrected for multiple comparisons with the false discovery rate (FDR, q values <0.05 (Benjamini & Hochberg, 1995)). Effect size were assessed using Wilcoxon effect size (r).

#### Association between neurobehavioural changes and microstructural changes after MBI

For each neurobehavioural and mean microstructural measures (i.e., FA, ICVF and ODI), score differences between assessment at pre-MBI and assessment at post-MBI were calculated for each participant. Score differences for neurobehavioural measures will be referred to as Δ neurobehavioural measures (neurobehavioural score post-MBI − neurobehavioural score pre-MBI). Score differences for mean FA, ICVF and ODI measures will be referred to as Δ mean FA, ICVF and ODI measures (i.e., mean FA post-MBI − mean FA pre-MBI; mean ICVF post-MBI – mean ICVF pre-MBI; mean ODI post-MBI − mean ODI pre-MBI). Negative Δ indicates a reduction of the scores between two time points, whereas positive Δ indicates an increase in scores between two time points.

Partial least square correlation analyses (PLSC) were performed to evaluate associations between changes in neurobehavioural measures and changes in mean microstructural measures after MBI. PLSC is a data-driven multivariate technique that maximizes the covariance between two matrices by identifying latent components (LCs) which are linear combinations of the two matrices, i.e., Δ neurobehavioural functioning measures and Δ mean microstructural measures (McIntosh & Lobaugh, 2004). A publicly available Matlab PLSC implementation was used: https://github.com/danizoeller/myPLS (Kebets et al., 2019; Zoller et al., 2019).

In a first PLSC, associations between changes in neurobehavioural measures and changes in mean FA measures after MBI were investigated to capture any changes related to white-matter microstructural properties measured by the classical tensor model.

The neurobehavioural functioning data refers to the three Δ neurobehavioural measures showing significant difference between the full-term and preterm group (i.e., BRIEF MI, BRIEF GEC, SDQ total), as well as age at assessment and gestational age at birth. The neurobehavioural functioning data were stored in a 32 × 5 matrix denoted X. Each row of X represents one subject and the matrix’s 5 columns are made up of the three Δ neurobehavioural measures showing significant difference between the full-term and preterm group, as well as age at assessment and gestational age at birth. The Δ mean FA data were gathered in a 32 × 50 matrix denoted Y, with each row matching one subject and each column one Δ mean FA measure for each of the 50 tracts extracted using the TractSeg framework. A cross-covariance matrix (which is effectively a correlation matrix, since the data are z-scores) was then computed between X and Y. Singular value decomposition was then applied to this cross-covariance matrix, resulting in latent components. Each latent component is composed of neurobehavioural functioning saliences and Δ mean FA saliences, and saliences indicate how strongly each neurobehavioural functioning measures and Δ mean FA measures contribute to the multivariate association of neurobehavioral functioning and Δ mean FA. The significance of latent components was determined by permutation testing (1000 permutations). Stability of neurobehavioural functioning saliences and Δ mean FA saliences were estimated using bootstrapping (500 bootstrap samples with replacement). Bootstrap ratio z-scores for each neurobehavioural functioning and Δ mean FA measures were obtained by dividing each neurobehavioural functioning and Δ mean FA salience by its bootstrap-estimated standard deviation, and a p-value was obtained for each bootstrap ratio z-score. Following the PLSC interpretation (Krishnan, Williams, McIntosh, & Abdi, 2011), the contribution of neurobehavioural functioning and Δ mean FA saliences for a given latent component was considered robust at p < 0.01 (i.e., absolute bootstrap ratio z-scores above 2.3 or below −2.3).

In a second PLSC, we investigated further associations between changes in neurobehavioural measures and changes in microstructural measures using the NODDI model, including mean ICVF and ODI measures after MBI. A procedure similar to the first PLSC was employed using a 32 × 5 matrix denoted X containing the neurobehavioural functioning data and a 32 × 100 matrix denoted Y containing the Δ mean ICVF and Δ mean ODI.

## Results

### Participant characteristics

The final sample included 32 preterm-born and 22 full-term young adolescents between 10 and 14 years of age. Baseline characteristics were similar between preterm and full-term participants for sex, age at the assessment and socio-economic status. The preterm group had significantly lower general ability index compared to the full-term group, see Table 1.

**Table 1.** Neonatal and demographic characteristics of the preterm and full-term participants

|  | **Preterm (n=32)** | **Full-term (n=22)** | **Group comparison** |
| --- | --- | --- | --- |
| **Birth weight**<br>mean (SD) in grams | 1240 (416.2) | 3403 (451.1) | $t(42.832) = -17.864, p < 0.001$ |
| **Gestational Age**<br>mean (SD) in weeks | 29.2 (1.9) | 39.8 (1.5) | $t(50.56) = -22.481, p < 0.001$ |
| **Age at assessment**<br>mean (SD) in years | 12.2 (1.2) | 11.9 (1.1) | $t(48.75) = 0.844, p = 0.403$ |
| **Sex, n** | 15 females<br>17 males | 9 females<br>13 males | $\chi^2(1, N = 55) = 0.188, p = 0.665$ |
| **Socio-economic status**<br>mean (SD) | 4 (2.7) | 3.1 (1.4) | $t(49.34) = 1.685, p = 0.098$ |
| **General ability index (GAI)**<br>mean (SD) | 109.3 (10.6) | 116.4 (11.4) | $t(43.25) = -2.288, p = 0.027$ |

\* Note: For preterm young adolescent, age correspond to the age at pre-MBI assessment. General ability index (GAI) was evaluated using the Wechsler Intelligence Scale for Children – 4th Edition, WISC-IV (Wechsler,
2003) as a measure of general intellectual functioning (mean for the general population = 100, standard-deviation = 15). Socioeconomic status of the parents was estimated using the Largo scale, a validated 12-point score based on maternal education and paternal occupation (Largo et al., 1989). Higher socio-economic status scores reflect lower socio-economic level. Independent-sample t-test or Chi-square as appropriate were used to compare the preterm and full-term participants. All p-values that survived false discovery rate (FDR) correction ( $q < 0.05$ ) are indicated in bold.

### Neurobehavioural outcomes: Comparison of the preterm and full-term groups

Neurobehavioural measures showing a significant difference and surviving FDR correction between the preterm and the full-term groups are presented in Figure 3. The preterm group showed higher BRIEF GEC (q < 0.001), BRIEF MI (q < 0.001), BRIEF BRI (q < 0.001) and SDQ total scores (q < 0.001) reflecting more executive function and behavioural difficulties in daily life compared to the full-term group. Moreover, the self-compassion score was reduced in the preterm group compared to the full-term group (q < 0.011), see Figure 3 and Supplementary Table S2. The other neurobehavioural scores were comparable between the preterm and full-term groups.

**Figure 3.**
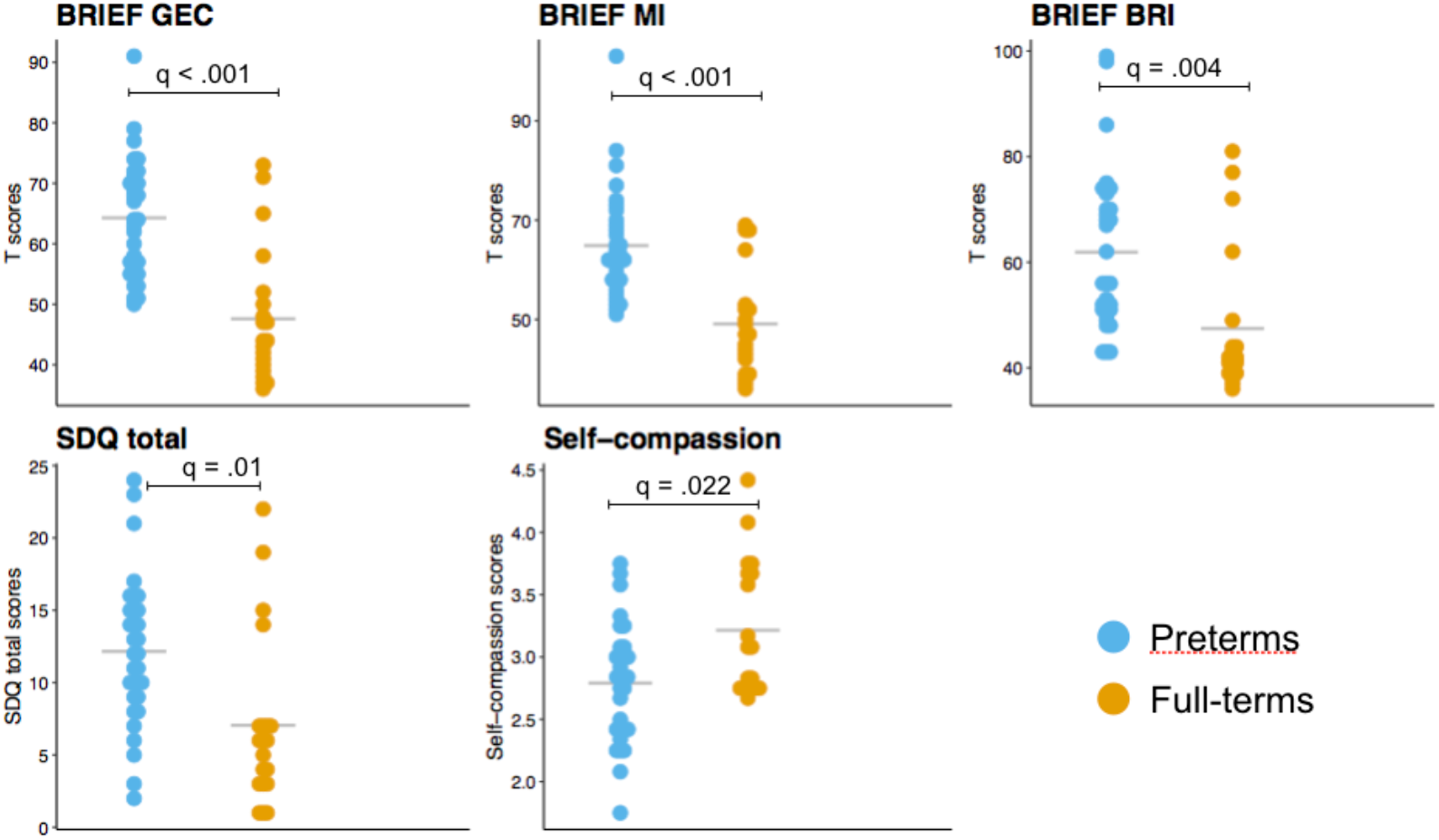
Neurobehavioural scores showing significant group differences between the preterm (prior to the MBI) and the full-term groups using independent sample t-test (FDR, q values <0.05). The mean for each measure and each group is represented by a grey line.

### Impact of MBI on neurobehavioral measures affected by preterm-birth

A significant decrease after MBI was observed for the following measures: BRIEF GEC (q < 0.00), BRIEF MI (q < 0.001) and SDQ total scores (q < 0.04), reflecting significantly less executive and behavioural difficulties in daily life. There was no significant difference before and after MBI for the BRIEF BRI and self-compassion scores, see Figure 4 and Supplementary Table S3.

**Figure 4.**
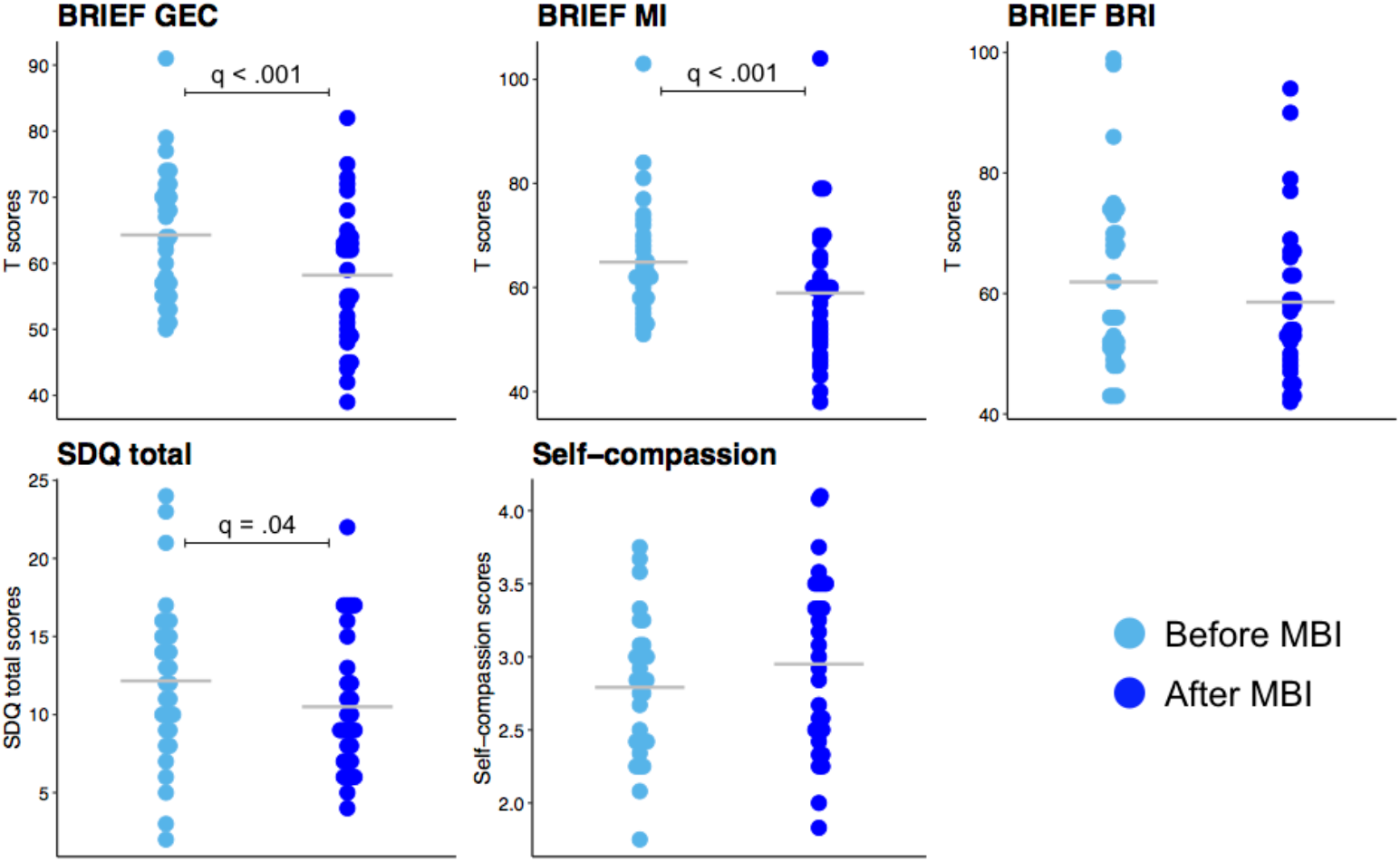
Neurobehavioural scores before and after MBI. Paired-sample t-tests was used to compared scores before and after MBI (FDR, q values <0.05). The mean for each measure and each group is represented by a grey line.

### Association between neurobehavioural changes and microstructural changes after MBI

The first PLSC analysis applied on neurobehavioural functioning (i.e., Δ neurobehavioural measures, age at the assessment and gestational age) and Δ mean FA identified one statistically significant latent component: latent component 1 (p = 0.005). Mean saliences as well as their bootstrap-estimated standard deviations for neurobehavioural functioning and Δ mean FA are reported in Supplementary Table S4. Latent component 1 revealed a general pattern of association between neurobehavioural functioning and Δ mean FA, see Figure 5. A decreased in Δ scores of the BRIEF GEC and BRIEF IM were associated with an increase in Δ mean FA values for a range of tracts, including the arcuate fascicle left/right, the anterior thalamic radiation left/right, the superior thalamic radiation right, the genu and isthmus of the corpus callosum, the cingulum left/right, the corticospinal tract left, the fronto-pontine tract left/right, the inferior cerebellar peduncle right, the inferior occipito-frontal fascicle left, the middle cerebellar peduncle, the optic radiation right, the superior cerebellar peduncle left/right, the superior longitudinal fascicle II and III left/right, the uncinate fascicle left, the thalamo-premotor and thalamo-occipital tract left/right, the striato-fronto-orbital left as well as the striato-premotor tract left/right. Moreover, lower gestational age was associated with a robust increased in Δ mean FA values in these same tracts, see Figure 5a. Importantly these microstructural changes were not associated with the age at the assessment of the participants.

**Figure 5.**
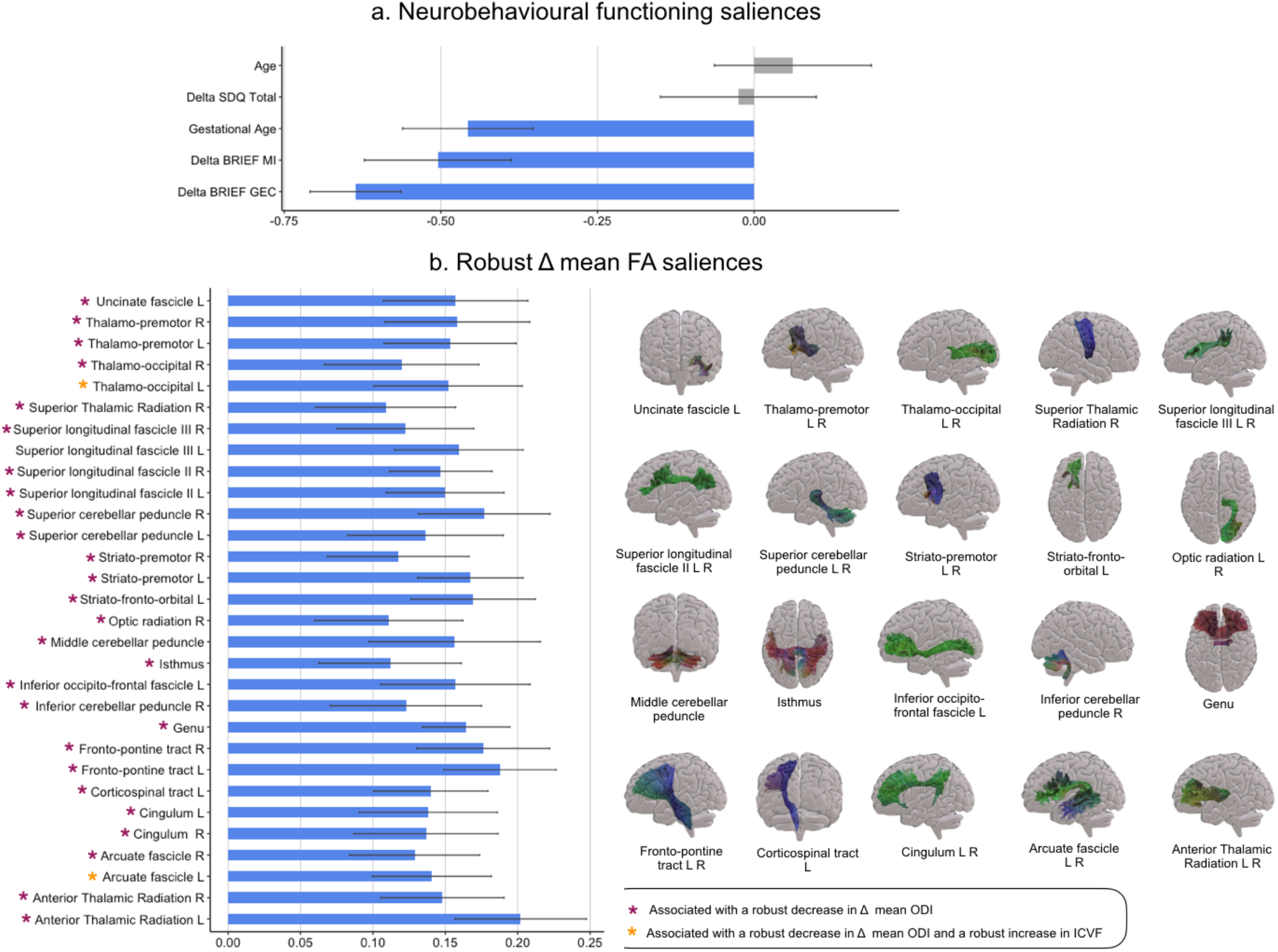
Associations between neurobehavioural functioning and Δ mean FA based on the PLSC analysis. A) Neurobehavioural functioning saliences: the diverging graph show mean saliences averaged across bootstrap samples and their bootstrap-estimated standard deviations (x-axis) for each neurobehavioural functioning measure (y-axis); robust saliences are represented in blue. Of note, saliences bellow 0 indicate a decreased in scores after MBI and saliences above 0 indicate an increased in scores after MBI. B) Robust Δ mean FA saliences: the diverging graph show robust mean saliences averaged across bootstrap samples and their bootstrap-estimated standard deviations (x-axis) for each Δ mean FA along a given tract (y-axis). The tracts extracted from TractSeg and showing robust Δ mean FA saliences are shown on the right side and the colour code for principal direction of diffusion is used: red (left – right), green (anterior – posterior), blue (inferior – superior). Based on the results from the second PLSC analyses: pink star indicates a robust negative contribution of Δ mean ODI along the given tract associated with a similar pattern of neurobehavioural functioning saliences; and orange star indicates both a robust negative contribution of Δ mean ODI and a robust positive contribution of Δ mean ICVF along the given tract associated with a similar pattern of neurobehavioural functioning saliences. Of note, saliences below 0 indicate a decreased in scores after MBI and saliences above 0 indicate an increased in scores after MBI; L left, R right.

The second PLSC analysis applied on neurobehavioural functioning (i.e., Δ neurobehavioural measures, age at the assessment and gestational age) and Δ mean ICVF and ODI identified one statistically significant latent component: latent component 1 (p = 0.02). Mean saliences as well as their bootstrap-estimated standard deviations for neurobehavioural functioning and Δ mean ICVF and ODI are reported in Supplementary Table S5 and Supplementary Figure S1. Latent component 1 revealed an association between decreased in Δ scores of the BRIEF GEC and BRIEF IM with a general decrease in Δ mean ODI for all tracts showing a robust increase in Δ mean FA, including: arcuate fascicle left/right, the anterior thalamic radiation left/right, the superior thalamic radiation right, the genu of the corpus callosum, the cingulum left/right, the corticospinal tract left, the fronto-pontine tract left/right, the inferior cerebellar peduncle right, the inferior occipito-frontal fascicle left, the middle cerebellar peduncle, the optic radiation right, the superior cerebellar peduncle left/right, the superior longitudinal fascicle II left/right and III right, the uncinate fascicle left, the thalamo-premotor and thalamo-occipital tract left/right, the striato-fronto-orbital left as well as the striato-premotor tract left/right; with the exception of the isthmus of the corpus callosum and the superior longitudinal fascicle III left, see Figure 5. This decreased in Δ scores of the BRIEF GEC and BRIEF IM were also associated with a combined increased in Δ mean ICVF and decrease in Δ mean ODI for left-lateralised tracts including: the arcuate fascicle, inferior cerebellar peduncle, thalamo-parietal and thalamo-occipital. Moreover, the decreased in Δ scores of the BRIEF GEC and BRIEF IM were associated with a robust reduction of ODI only on the rostrum of the corpus callosum, on the superior longitudinal fascicle I left and on the striato-fronto-occipital right. Finally, the general increase in Δ ICVF and decrease in ODI measures were also associated with higher gestational age in VPT young adolescents. Importantly these microstructural changes were not associated with the age of the participants.

## Discussion

In this study, we investigated the benefits of an 8-weeks MBI on neurobehavioral functioning and its association with white-matter microstructural changes in VPT young adolescents using both DTI and NODDI parameters. Our finding showed an enhancement of global executive functioning in daily life after MBI that was associated with a general pattern of significant increase in FA along with a decrease in ODI in a range of white-matter tracts involved in executive processes. This general pattern of increase FA and decrease ODI values was also negatively associated with gestational age, meaning that the microstructural changes in FA and ODI after MBI was particularly marked in young adolescents with lower gestational age.

In the present study, preterm young adolescents showed significant difficulties in executive and behavioural functioning in daily life as measured by the BRIEF and SDQ parent questionnaires, as well as significantly lower self-compassion using a self-reported questionnaire compared to full-term controls. These weaknesses have been previously described in several cohorts of children, adolescents and adults born prematurely (Aarnoudse-Moens, Weisglas-Kuperus, van Goudoever, & Oosterlaan, 2009; Luu, Ment, Allan, Schneider, & Vohr, 2011; Mulder, Pitchford, Hagger, & Marlow, 2009). After an 8-weeks MBI, an enhancement of executive and behavioural functioning in daily life of preterm young adolescents was observed. These behavioural results corroborate the results from our previous study using a gold standard randomised controlled trial design (Siffredi, Liverani, Hüppi, et al., 2021), which indicates that the gain observed after MBI on these executive and behavioural measures is independent of test-retest effect. The benefits of MBI observed on daily executive and behavioural functioning are also consistent with previous studies completed in other populations (Flook, Goldberg, Pinger, & Davidson, 2015; Flook et al., 2010; Geronimi et al., 2019; van de Weijer-Bergsma, Formsma, de Bruin, & Bogels, 2012; van der Oord, Bogels, & Peijnenburg, 2012).

Combining both the classical DTI model and the NODDI model (i.e., a multi-component model that captures neurite morphology), microstructural changes associated with the beneficial effect of MBI on executive and behavioural functioning were explored. Our finding show that the gain observed in executive functioning after an 8-week MBI was associated with a general pattern of increase in FA along with a decrease in ODI. While an increase in FA values reflect changes in the diffusion of water molecules along a given tract, the decrease in axonal dispersion indicate more specifically an enhancement in the coherence of the spatial organisation of the axons (Mah, Geeraert, & Lebel, 2017). This pattern of higher FA coupled with decrease ODI in a specific white-matter tract is commonly interpreted as an indication of increased connectivity (Laneri et al., 2016). Importantly, this pattern of microstructural changes associated with improved executive functioning after MBI was observed in white-matter tracts known to be involved in different executive processes, i.e., attentional control, inhibition, processing speed, updating, fluency and self-regulation (Bettcher et al., 2016; Diao et al., 2015; Frye et al., 2010; Mamiya, Richards, & Kuhl, 2018) or in white-matter tracts implicated in sensorimotor circuits that support executive functioning (Koziol & Lutz, 2013), including the anterior and superior thalamic radiation, the genu of the corpus callosum, the cingulum, the fronto-pontine tract, the inferior occipito-frontal fascicle, the superior longitudinal fascicle II and III, the arcuate fascicle, the thalamo-premotor and thalamo-occipital tract, the striato-fronto-orbital as well as the striato-premotor tract. Moreover, the uncinate fascicle, known to be involved in emotional-regulation processes, also showed this pattern of increase FA and reduced ODI associated with improvement in executive functioning after MBI (Oishi et al., 2015; Sobhani, Baker, Martins, Tuvblad, & Aziz-Zadeh, 2015). These results corroborate previous research studies showing that the practice of MBI or MBI-related approaches such as meditation are associated with increases in FA in these tracts or related-regions of interest (Hölzel et al., 2016; Kang et al., 2013; Laneri et al., 2016; Luders et al., 2011; Luders et al., 2012; Patsenko et al., 2019; Tang et al., 2012). Moreover, there was an association between changes in executive functioning after MBI with changes in cerebellar mean FA including, the inferior cerebellar peduncle, the middle cerebellar peduncle and the superior cerebellar peduncles. Again, these FA changes were more specifically coupled with a decrease in neurite orientation dispersion. Cerebellar regions have been implicated in executive functions and proposed to play a role of “control of behaviour” by linking movement to though (Berquin et al., 1998; Koziol, Budding, & Chidekel, 2012; Schweizer et al., 2008). This is also in line with previous results showing changes in grey matter density of cerebellar regions after an 8-week Mindfulness program (Hölzel et al., 2011). Compared to previous findings, the use of the NODDI model in the current study allows to characterise more precisely this increase in FA explained by a concurrent decrease in neurite orientation dispersion.

Altogether, the enhancement in executive functioning was not only associated with FA changes in white-matter tracts involved in executive functioning per se; but it was also associated with FA changes in white-matter tracts implicated in bottom-up and sensorimotor interaction on which executive functions rely. Importantly, the NODDI model allowed to characterise more precisely this general increase in FA with concurrent decrease in neurite orientation dispersion. As opposed to executive functioning, the changes observed in socio-emotional and behavioural functioning after MBI, as measured by the SDQ total score, were not significantly associated with changes in mean FA.

Additionally, the increase in FA and reduction in ODI values following MBI of these tracts was associated with lower gestational age at birth. In other words, the microstructural gain observed on the tracts mentioned above was bigger in the most preterm young adolescents. It is likely that MBI is most beneficial for the most vulnerable young adolescents as they have more opportunities for further enhancements. This is consistent with the results of our randomised controlled trial showing individual variability in the response to MBI with a greater benefit on overall executive skills in VPT young adolescents born with higher risk (i.e., smaller gestational age and lower weight at birth), in comparison with VPT young adolescents born with lower risk (Siffredi, Liverani, Hüppi, et al., 2021). This pattern of increased benefit of MBI in the most vulnerable individuals has already been observed in children with weaker executive abilities (Flook et al., 2010).

Finally, the reduction in executive difficulties observed after an 8-week MBI was also associated with an increase in neurite density (ICVF) in white-matter tracts implicated in executive functioning, including the arcuate fasciculus as well as thalamic and cerebellar tracts. This pattern reflecting an increase in packing density of axons in white matter in a few tracts was also associated with reduced gestational age at birth. These results go in line with the suggested increased benefit of MBI in the most vulnerable individuals and in individuals with weaker executive abilities (Flook et al., 2010; Siffredi, Liverani, Hüppi, et al., 2021).

Our findings should also be considered in light of the limitations of the current study. The beneficial effect of MBI on executive and socio-emotional competences were observed through parent-questionnaires. These changes observed via parents-reported questionnaires might be questionable as parents were not blind to the intervention. This point might be methodologically improved via the inclusion of an active control condition or placebo condition, as done in previous studies (Britton et al., 2014; Pascoe et al., 2013). Nevertheless, the association found with micro-structural changes on specific tracks suggest that the benefit of MBI observed via parent questionnaire are well-grounded.

## Conclusions

This study provides evidence that the enhancement in executive functioning after an MBI in VPT young adolescents is associated with white-matter microstructural changes in tracts involved in executive processes as well as in tracts involved in sensorimotor circuit supporting and interacting with executive processes. Our study also suggests that the most vulnerable VPT young adolescents, i.e., the ones with lowest gestational age at birth, show the biggest gain in terms of white-matter microstructural changes. Finally, MBI appears to be a promising tool for enhancing executive functioning and white-matter brain plasticity in a vulnerable population such as VPT young adolescents.

## Data Availability

Ethical restrictions prevent us from making anonymised data available in a public repository. Data may be available for researchers to researchers who meet the criteria for access to confidential data by direct request to the "Mindful preterm teens study" team. There are restrictions on data related to identifying participant information and appropriate ethical approval is required prior to release. Only de-identified data will be available.

## Abbreviations

NEPSY-II: A Developmental Neuropsychological Assessment, 2nd Edition
AD: Axial Diffusivity
BRIEF: Behaviour Rating Inventory of Executive Function-parent version
BRI: Behavioural Regulation Index
DTI: Diffusion Tensor Imaging
DW: Diffusion-Weighted
FDR: False Discovery Rate
FOD: Fibre Orientation Distributions
fODF: Fibre Orientation Distribution Function
FA: Fractional Anisotropy
GAI: General Ability Index
GEC: Global Executive Composite
ICVF: Intra-cellular Volume Fraction
LCs: Latent Components
MRI: Magnetic Resonance Imaging
MI: Mean Diffusivity
MD: Metacognition Index
MBCT: Mindfulness-Based Cognitive Therapy
MBI: Mindfulness-Based Intervention
MBSR: Mindfulness-Based Stress Reduction
MSMT-CSD: Multi-Shell Multi-Tissue Constrained Spherical Deconvolution
NODDI: Neurite Orientation Dispersion and Density Imaging
(ODI): Orientation Dispersion Index
PLSC: Partial Least Square Correlation Analyses
RD: Radial Diffusivity
SDQ: Strength and Difficulties Questionnaire
TCC: Temporal Context Confusion index
TractSeg: Tractography-Based Segmentation
VPT: Very Preterm
WISC-IV: Wechsler Intelligence Scale for Children, 4th Edition

## Acknowledgements

We thank and acknowledge all participating young adolescents and families who made this research possible. We also thank the Fondation Campus Biotech Geneva (FCBG), a foundation of the Swiss Federal Institute of Technology Lausanne (EPFL), the University of Geneva (UniGe), and the University Hospitals of Geneva (HUG); the Research Platform of the University Hospitals of Geneva (HUG) for their practical help; as well as Mariana Magnus Smith and Françoise Stuckelberger-Grobéty for their implication as MBI instructors.

## References

Aarnoudse-Moens, C. S., Weisglas-Kuperus, N., van Goudoever, J. B., & Oosterlaan, J. (2009). Meta-analysis of neurobehavioral outcomes in very preterm and/or very low birth weight children. Pediatrics, 124(2), 717–728. doi:10.1542/peds.2008-2816

Allin, M. P., Kontis, D., Walshe, M., Wyatt, J., Barker, G. J., Kanaan, R. A., … Nosarti, C. (2011). White matter and cognition in adults who were born preterm. PLoS One, 6(10), e24525.

Anderson, P. (2002). Assessment and development of executive function (EF) during childhood. Child Neuropsychol, 8(2), 71–82. doi:10.1076/chin.8.2.71.8724

Barnes, V. A., Bauza, L. B., & Treiber, F. A. (2003). Impact of stress reduction on negative school behavior in adolescents. Health Qual Life Outcomes, 1, 10.

Beauchemin, J., Hutchins, T. L., & Patterson, F. (2008). Mindfulness meditation may lessen anxiety, promote social skills, and improve academic performance among adolescents with learning disabilities. Complement Health Pract Rev, 13(1), 34–45.

Beaulieu, C. (2002). The basis of anisotropic water diffusion in the nervous system - a technical review. NMR Biomed, 15(7-8), 435–455. doi:10.1002/nbm.782

Beaulieu, C. (2002). The basis of anisotropic water diffusion in the nervous system–a technical review. NMR in Biomedicine: An International Journal Devoted to the Development Application of Magnetic Resonance In Vivo, 15(7-8), 435–455.

Benjamini, Y., & Hochberg, Y. (1995). Controlling the false discovery rate: a practical and powerful approach to multiple testing. Journal of the Royal Statistical Society, 57, 289–300.

Berquin, P., Giedd, J., Jacobsen, L., Hamburger, S., Krain, A., Rapoport, J., & Castellanos, F. (1998). Cerebellum in attention-deficit hyperactivity disorder: a morphometric MRI study. Neurology, 50(4), 1087–1093.

Best, J. R., Miller, P. H., & Naglieri, J. A. (2011). Relations between Executive Function and Academic Achievement from Ages 5 to 17 in a Large, Representative National Sample. Learn Individ Differ, 21(4), 327–336. doi:10.1016/j.lindif.2011.01.007

Bettcher, B. M., Mungas, D., Patel, N., Elofson, J., Dutt, S., Wynn, M., … Kramer, J. H. (2016). Neuroanatomical substrates of executive functions: beyond prefrontal structures. Neuropsychologia, 85, 100–109.

Biegel, G. M., Brown, K. W., Shapiro, S. L., & Schubert, C. M. (2009). Mindfulness-based stress reduction for the treatment of adolescent psychiatric outpatients: A randomized clinical trial. J Consult Clin Psychol, 77(5), 855–866. doi:10.1037/a0016241

Black, D. S., Milam, J., & Sussman, S. (2009). Sitting-meditation interventions among youth: a review of treatment efficacy. Pediatrics, 124(3), e532–541. doi:10.1542/peds.2008-3434

Boorman, E. D., O’Shea, J., Sebastian, C., Rushworth, M. F., & Johansen-Berg, H. (2007). Individual differences in white-matter microstructure reflect variation in functional connectivity during choice. Current Biology, 17(16), 1426–1431.

Britton, W. B., Lepp, N. E., Niles, H. F., Rocha, T., Fisher, N. E., & Gold, J. S. (2014). A randomized controlled pilot trial of classroom-based mindfulness meditation compared to an active control condition in sixth-grade children. J Sch Psychol, 52(3), 263–278. doi:10.1016/j.jsp.2014.03.002

Broderick, P. C., & Metz, S. (2009). Learning to BREATHE: A pilot trial of a mindfulness curriculum for adolescents. Advances in School Mental Health Promotion, 2, 35–46

Carter, A. S., Briggs-Gowan, M. J., & Davis, N. O. (2004). Assessment of young children’s social-emotional development and psychopathology: recent advances and recommendations for practice. J Child Psychol Psychiatry, 45(1), 109–134. doi:10.1046/j.0021-9630.2003.00316.x

Chandio, B., Harezlak, J., & Garyfallidis, E. (2019). Bundle Analytics: a computational and statistical analysis framework for tractometric studies. Paper presented at the Organization for Human Brain Mapping.

Christ, S. E., Kester, L. E., Bodner, K. E., & Miles, J. H. (2011). Evidence for selective inhibitory impairment in individuals with autism spectrum disorder. Neuropsychology, 25(6), 690–701. doi:10.1037/a0024256

Cordero-Grande, L., Christiaens, D., Hutter, J., Price, A. N., & Hajnal, J. V. J. N. (2019). Complex diffusion-weighted image estimation via matrix recovery under general noise models. 200, 391–404.

de Bie, H. M., Boersma, M., Wattjes, M. P., Adriaanse, S., Vermeulen, R. J., Oostrom, K. J., … Delemarre-Van de Waal, H. A. J. E. j. o. p. (2010). Preparing children with a mock scanner training protocol results in high quality structural and functional MRI scans. 169(9), 1079–1085.

De Vos, T. (1994). Tempo Test Rekenen [Tempo Test Arithmetic]. Handleiding Tempo-Test-Rekenen (2nd edition). Lisse, The Netherlands: Swets Test Publishers.

Diao, L., Yu, H., Zheng, J., Chen, Z., Huang, D., & Yu, L. (2015). Abnormalities of the uncinate fasciculus correlate with executive dysfunction in patients with left temporal lobe epilepsy. Magnetic Resonance Imaging, 33(5), 544–550.

Felver, J. C., Tipsord, J. M., Morris, M. J., Racer, K. H., & Dishion, T. J. (2017). The Effects of Mindfulness-Based Intervention on Children’s Attention Regulation. 21(10), 872–881. doi:10.1177/1087054714548032

Flook, L., Goldberg, S. B., Pinger, L., & Davidson, R. J. (2015). Promoting prosocial behavior and self-regulatory skills in preschool children through a mindfulness-based Kindness Curriculum. Dev Psychol, 51(1), 44–51. doi:10.1037/a0038256

Flook, L., Smalley, S. L., Kitil, M. J., Galla, B. M., Kaiser-Greenland, S., Locke, J., … Kasari, C. (2010). Effects of Mindful Awareness Practices on Executive Functions in Elementary School Children. Journal of Applied School Psychology, 26(1), 70–95. doi:10.1080/15377900903379125

Frye, R. E., Hasan, K., Malmberg, B., Desouza, L., Swank, P., Smith, K., & Landry, S. (2010). Superior longitudinal fasciculus and cognitive dysfunction in adolescents born preterm and at term. Developmental Medicine Child Neurology, 52(8), 760–766.

Geronimi, E. M. C., Arellano, B., & Woodruff-Borden, J. (2019). Relating mindfulness and executive function in children. Clin Child Psychol Psychiatry, 1359104519833737. doi:10.1177/1359104519833737

Gioia, G., Isquith, P., Guy, S., & Kenworthy, L. (2000). BRIEF – Behavior Rating Inventory of Executive Function. Professional manual. Odessa, FL: Psychological Assessment Resources Inc.

Goodman, R. (1997). The Strengths and Difficulties Questionnaire: a research note. J Child Psychol Psychiatry, 38(5), 581–586.

Goodman, R. (2001). Psychometric properties of the strengths and difficulties questionnaire. J Am Acad Child Adolesc Psychiatry, 40(11), 1337–1345. doi:10.1097/00004583-200111000-00015

Holmes, J., & Adams, J. W. (2006). Working Memory and Children’s Mathematical Skills: Implications for mathematical development and mathematics curricula. Educational Psychology, 26(3). doi:10.1080/01443410500341056

Hölzel, B. K., Brunsch, V., Gard, T., Greve, D. N., Koch, K., Sorg, C., … Milad, M. R. (2016). Mindfulness-based stress reduction, fear conditioning, and the uncinate fasciculus: a pilot study. Frontiers in behavioral neuroscience, 10, 124.

Hölzel, B. K., Carmody, J., Vangel, M., Congleton, C., Yerramsetti, S. M., Gard, T., & Lazar, S. W. (2011). Mindfulness practice leads to increases in regional brain gray matter density. Psychiatry research: neuroimaging, 191(1), 36–43.

Izard, C. E. (2011). Forms and functions of emotions: Matters of emotion–cognition interactions. Emotion Review, 3, 371–378.

Jenkinson, M., Beckmann, C. F., Behrens, T. E., Woolrich, M. W., & Smith, S. M. J. N. (2012). Fsl. 62(2), 782–790.

Jeurissen, B., Tournier, J.-D., Dhollander, T., Connelly, A., & Sijbers, J. J. N. (2014). Multi-tissue constrained spherical deconvolution for improved analysis of multi-shell diffusion MRI data. 103, 411–426.

Jones, D. K., Knösche, T. R., & Turner, R. (2013). White matter integrity, fiber count, and other fallacies: the do’s and don’ts of diffusion MRI. Neuroimage, 73, 239–254.

Jones, K. M., Champion, P. R., & Woodward, L. J. (2013). Social competence of preschool children born very preterm. Early Hum Dev, 89(10), 795–802.

Kabat-Zinn, J. (2003). Mindfulness-Based interventions in context: Past, present and future. Clinical Psychology: Science and Practice, 10, 144–156. doi:DOI: 10.1093/clipsy.bpg016

Kang, D. H., Jo, H. J., Jung, W. H., Kim, S. H., Jung, Y. H., Choi, C. H., … Kwon, J. S. (2013). The effect of meditation on brain structure: cortical thickness mapping and diffusion tensor imaging. Soc Cogn Affect Neurosci, 8(1), 27–33. doi:10.1093/scan/nss056

Kebets, V., Holmes, A. J., Orban, C., Tang, S., Li, J., Sun, N., … Yeo, B. T. T. (2019). Somatosensory-Motor Dysconnectivity Spans Multiple Transdiagnostic Dimensions of Psychopathology. Biological Psychiatry, 0(0).

Kellner, E., Dhital, B., Kiselev, V. G., & Reisert, M. J. M. r. i. m. (2016). Gibbs-ringing artifact removal based on local subvoxel-shifts. 76(5), 1574–1581.

Korkman, M., Kirk, U., & Kemp, S. (2007). A developmental neuropsychological assessment 2nd Edition: NEPSY-II. San Antonio, Texas: Pearson.

Koziol, L. F., Budding, D. E., & Chidekel, D. (2012). From movement to thought: executive function, embodied cognition, and the cerebellum. The Cerebellum, 11(2), 505–525.

Koziol, L. F., & Lutz, J. T. (2013). From movement to thought: the development of executive function. Applied Neuropsychology: Child, 2(2), 104–115.

Krishnan, A., Williams, L. J., McIntosh, A. R., & Abdi, H. (2011). Partial Least Squares (PLS) methods for neuroimaging: a tutorial and review. Neuroimage, 56(2), 455–475. doi:10.1016/j.neuroimage.2010.07.034

Laneri, D., Schuster, V., Dietsche, B., Jansen, A., Ott, U., & Sommer, J. (2016). Effects of long-term mindfulness meditation on brain’s white matter microstructure and its aging. Frontiers in aging neuroscience, 7, 254.

Largo, R. H., Pfister, D., Molinari, L., Kundu, S., Lipp, A., & Duc, G. (1989). Significance of prenatal, perinatal and postnatal factors in the development of AGA preterm infants at five to seven years. Dev Med Child Neurol, 31(4), 440–456.

Lee, J., Semple, R. J., Rosa, D., & Miller, L. (2008). Mindfulness-based cognitive therapy for children: results of a pilot study. J Cogn Psychother, 22(1), 15–28.

Liverani, M. C., Freitas, L. G. A., Siffredi, V., Mikneviciute, G., Martuzzi, R., Meskaldij, D. E., Hüppi, P. S. (2020). Get real: Orbitofrontal cortex mediates the ability to sense reality in early adolescents. Brain and Behavior. doi:https://doi.org/10.1002/brb3.1552

Liverani, M. C., Manuel, A. L., Nahum, L., Guardabassi, V., Tomasetto, C., & Schnider, A. (2017). Children’s sense of reality: The development of orbitofrontal reality filtering. Child Neuropsychology, 23(4), 408–421. doi:10.1080/09297049.2015.1120861

Loe, I. M., Lee, E. S., & Feldman, H. M. (2013). Attention and internalizing behaviors in relation to white matter in children born preterm. Journal of developmental behavioral pediatrics, 34(3), 156.

Luders, E., Clark, K., Narr, K. L., & Toga, A. W. (2011). Enhanced brain connectivity in long-term meditation practitioners. Neuroimage, 57(4), 1308–1316. doi:10.1016/j.neuroimage.2011.05.075

Luders, E., Phillips, O. R., Clark, K., Kurth, F., Toga, A. W., & Narr, K. L. (2012). Bridging the hemispheres in meditation: thicker callosal regions and enhanced fractional anisotropy (FA) in long-term practitioners. Neuroimage, 61(1), 181–187. doi:10.1016/j.neuroimage.2012.02.026

Luu, T. M., Ment, L., Allan, W., Schneider, K., & Vohr, B. R. (2011). Executive and memory function in adolescents born very preterm. Pediatrics, 127(3), e639–646. doi:10.1542/peds.2010-1421

Mah, A., Geeraert, B., & Lebel, C. (2017). Detailing neuroanatomical development in late childhood and early adolescence using NODDI. PLoS One, 12(8), e0182340.

Mamiya, P. C., Richards, T. L., & Kuhl, P. K. (2018). Right forceps minor and anterior thalamic radiation predict executive function skills in young bilingual adults. Frontiers in psychology, 9, 118.

Marchand, W. R. (2014). Neural mechanisms of mindfulness and meditation: Evidence from neuroimaging studies. World Journal of Radiology, 6(7), 471–479. doi:10.4329/wjr.v6.i7.471

McIntosh, A. R., & Lobaugh, N. J. J. N. (2004). Partial least squares analysis of neuroimaging data: applications and advances. 23, S250–S263.

Mikolajczak, M., Quoidbach, J., Kotsou, I., & Nelis, D. (2009). Les compétences émotionnelles. Paris, France: Dunod.

Montagna, A., & Nosarti, C. (2016). Socio-emotional development following very preterm birth: pathways to psychopathology. Frontiers in psychology, 7, 80.

Mulder, H., Pitchford, N. J., Hagger, M. S., & Marlow, N. (2009). Development of Executive Function and Attention in Preterm Children: A Systematic Review. Developmental Neuropsychology, 34(4), 393–421. doi:10.1080/87565640902964524

Napoli, M., Krech, P. R., & Holley, L. C. (2004). Mindfulness training for elementary school students: the attention academy. J Appl Sch Psychol, 21(1), 99–123.

Nosarti, C., Allin, M. P., Frangou, S., Rifkin, L., & Murray, R. M. (2005). Hyperactivity in adolescents born very preterm is associated with decreased caudate volume. Biol Psychiatry, 57(6), 661–666.

Oishi, K., Faria, A. V., Hsu, J., Tippett, D., Mori, S., & Hillis, A. E. (2015). Critical role of the right uncinate fasciculus in emotional empathy. Annals of Neurology, 77(1), 68–74.

Pascoe, L., Roberts, G., Doyle, L. W., Lee, K. J., Thompson, D. K., Seal, M. L., … Anderson, P. J. (2013). Preventing academic difficulties in preterm children: a randomised controlled trial of an adaptive working memory training intervention - IMPRINT study. BMC Pediatr, 13, 144. doi:10.1186/1471-2431-13-144

Patsenko, E. G., Adluru, N., Birn, R. M., Stodola, D. E., Kral, T. R., Farajian, R., … Davidson, R. J. (2019). Mindfulness video game improves connectivity of the fronto-parietal attentional network in adolescents: A multi-modal imaging study. Scientific reports, 9(1), 1–8.

Perlman, J. M. (1998). White matter injury in the preterm infant: an important determination of abnormal neurodevelopment outcome. Early Hum Dev, 53(2), 99–120.

Pines, A. R., Cieslak, M., Larsen, B., Baum, G. L., Cook, P. A., Adebimpe, A., … Murtha, K. (2020). Leveraging multi-shell diffusion for studies of brain development in youth and young adulthood. Developmental cognitive neuroscience, 43, 100788.

Raes, F., Pommier, E., Neff, K. D., & Van Gucht, D. (2011). Construction and factorial validation of a short form of the Self-Compassion Scale. Clin Psychol Psychother, 18(3), 250–255. doi:10.1002/cpp.702

Robitail, S., Ravens-Sieberer, U., Simeoni, M. C., Rajmil, L., Bruil, J., Power, M., … Group, K. (2007). Testing the structural and cross-cultural validity of the KIDSCREEN-27 quality of life questionnaire. Qual Life Res, 16(8), 1335–1345. doi:10.1007/s11136-007-9241-1

Schonert-Reichl, K. A., & Lawlor, M. S. (2010). The effects of a mindfulness-based education program on pre-and early adolescents’ well-being and social and emotional competence. Mindfulness, 1, 137–151. doi:10.1007/s12671-010-0011-8

Schonert-Reichl, K. A., Oberle, E., Lawlor, M. S., Abbott, D., Thomson, K., Oberlander, T. F., & Diamond, A. (2015). Enhancing cognitive and social-emotional development through a simple-to-administer mindfulness-based school program for elementary school children: a randomized controlled trial. Dev Psychol, 51(1), 52–66. doi:10.1037/a0038454

Schweizer, T. A., Levine, B., Rewilak, D., O’Connor, C., Turner, G., Alexander, M. P., … Stuss, D. T. (2008). Rehabilitation of executive functioning after focal damage to the cerebellum. Neurorehabilitation Neural Repair, 22(1), 72–77.

Segal, Z. V., Williams, J. M. G., & Teasdale, J. D. (2001). Mindfulness-based cognitive theraphy for depression: a new approach to preventing relapse: Guilford Publications.

Semple, R. J., Lee, J., Rosa, D., & Miller, L. (2009). A Randomized Trial of Mindfulness-Based Cognitive Therapy for Children: Promoting Mindful Attention to Enhance Social-Emotional Resiliency in Children. Journal of Child and Family Studies, 19(2), 218–229. doi:DOI: 10.1007/s10826-009-9301-y

Sharp, P. B., Sutton, B. P., Paul, E. J., Sherepa, N., Hillman, C. H., Cohen, N. J., … Telzer, E. H. (2018). Mindfulness training induces structural connectome changes in insula networks. Scientific reports, 8(1), 7929.

Siffredi, V., Liverani, M. C., Hüppi, P. S., Freitas, L., De Albuquerque, J., Gimbert, F., … Ha-Vinh Leuchter, R. (2021). Mindfulness-based intervention for very preterm young adolescents: An RCT. MedRxiv: https://www.medrxiv.org/content/10.1101/2021.01.19.21250087v1, 2021.2003.2014.21253449. doi:10.1101/2021.03.14.21253449

Siffredi, V., Liverani, M. C., Magnus-Smith, M., Meskaldji, D., Stuckelberger-Grobety, F., Freitas, L., … Ha-Vinh Leuchter, R. (2021). Improving executive, behavioural and socio-emotional competences in very preterm young adolescents through a mindfulness-based intervention: study protocol and feasibility. MedRxiv: https://www.medrxiv.org/content/10.1101/2021.01.19.21250087v1.

Smith, S. M. J. H. b. m. (2002). Fast robust automated brain extraction. 17(3), 143–155.

Sobhani, M., Baker, L., Martins, B., Tuvblad, C., & Aziz-Zadeh, L. (2015). Psychopathic traits modulate microstructural integrity of right uncinate fasciculus in a community population. NeuroImage: Clinical, 8, 32–38.

Soria-Pastor, S., Gimenez, M., Narberhaus, A., Falcon, C., Botet, F., Bargallo, N., … Junque, C. (2008). Patterns of cerebral white matter damage and cognitive impairment in adolescents born very preterm. International Journal of Developmental Neuroscience, 26(7), 647–654.

Tamnes, C. K., Roalf, D. R., Goddings, A.-L., & Lebel, C. (2018). Diffusion MRI of white matter microstructure development in childhood and adolescence: Methods, challenges and progress. Developmental cognitive neuroscience, 33, 161–175.

Tang, Y. Y., Lu, Q., Fan, M., Yang, Y., & Posner, M. I. (2012). Mechanisms of white matter changes induced by meditation. Proc Natl Acad Sci U S A, 109(26), 10570–10574. doi:10.1073/pnas.1207817109

Team, R. (Producer). (2020). RStudio: Integrated Development for R.

Team, R. C. (Producer). (2019). R: A language and environment for statistical computing.

Thompson, D. K., Lee, K. J., Egan, G. F., Warfield, S. K., Doyle, L. W., Anderson, P. J., & Inder, T. E. (2014). Regional white matter microstructure in very preterm infants: predictors and 7 year outcomes. Cortex, 52, 60–74.

Tournier, J.-D., Smith, R., Raffelt, D., Tabbara, R., Dhollander, T., Pietsch, M., … Connelly, A. J. N. (2019). MRtrix3: A fast, flexible and open software framework for medical image processing and visualisation. 202, 116137.

van de Weijer-Bergsma, E., Formsma, A. R., de Bruin, E. I., & Bogels, S. M. (2012). The Effectiveness of Mindfulness Training on Behavioral Problems and Attentional Functioning in Adolescents with ADHD. J Child Fam Stud, 21(5), 775–787. doi:10.1007/s10826-011-9531-7

van der Oord, S., Bogels, S. M., & Peijnenburg, D. (2012). The Effectiveness of Mindfulness Training for Children with ADHD and Mindful Parenting for their Parents. J Child Fam Stud, 21(1), 139–147. doi:10.1007/s10826-011-9457-0

Vaughan, L., & Giovanello, K. (2010). Executive function in daily life: Age-related influences of executive processes on instrumental activities of daily living. Psychol Aging, 25(2), 343–355. doi:10.1037/a0017729

Veraart, J., Fieremans, E., & Novikov, D. S. J. M. r. i. m. (2016). Diffusion MRI noise mapping using random matrix theory. 76(5), 1582–1593.

Veraart, J., Novikov, D. S., Christiaens, D., Ades-Aron, B., Sijbers, J., & Fieremans, E. J. N. (2016). Denoising of diffusion MRI using random matrix theory. 142, 394–406.

Visu-Petra, L., Cheie, L., Benga, O., & Miclea, M. (2011). Cognitive control goes to school: The impact of executive functions on academic performance. Social and Behavioral Sciences, 11, 240–244.

Volpe, J. (2003). Cerebral white matter injury of the premature infant—more common than you think. Pediatrics, 112(1), 176–180.

Wasserthal, J., Neher, P., & Maier-Hein, K. H. J. N. (2018). TractSeg-Fast and accurate white matter tract segmentation. 183, 239–253.

Wechsler, D. (2003). Manual for the Wechsler Intelligence Scale for Children-IV. New York: Psychological Corporation.

Wentzel, K. R. (1994). Relations of social goal pursuit to social acceptance, classroom behavior, and perceived social support. Journal of Educational Psychology, 86(2), 173–182.

Woodward, L. J., Clark, C. A., Pritchard, V. E., Anderson, P. J., & Inder, T. E. (2011). Neonatal white matter abnormalities predict global executive function impairment in children born very preterm. Developmental Neuropsychology, 36(1), 22–41.

Zhang, H., Schneider, T., Wheeler-Kingshott, C. A., & Alexander, D. C. (2012). NODDI: practical in vivo neurite orientation dispersion and density imaging of the human brain. Neuroimage, 61(4), 1000–1016.

Zoller, D., Sandini, C., Karahanoglu, F. I., Padula, M. C., Schaer, M., Eliez, S., & Van De Ville, D. (2019). Large-Scale Brain Network Dynamics Provide a Measure of Psychosis and Anxiety in 22q11.2 Deletion Syndrome. Biological Psychiatry: Cognitive Neuroscience and Neuroimaging, 4, 881–892 doi:10.1016/j.bpsc.2019.04.004

