## Supplementary Materials for "Mindfulness-based intervention in preterm young adolescents: benefits on neurobehavioural functioning and its association with white-matter microstructural changes"

#### Supplementary Tables

**Supplementary Table S1**. Details of the neurobehavioural measures and scores

| Domains | Modalities | Measures | Description | Scores |
|---|---|---|---|---|
| **Executive competences** | | | | |
| | Parent questionnaire | | | |
| | | Behaviour Rating Inventory of Executive Function, parent version (BRIEF, Gioia, Isquith, Guy, and Kenworthy (2000)) | | |
| | | | The BRIEF parent questionnaire provides an index of attention, hyperactivity and impulsivity in everyday life. The BRIEF comprises 86 items over two standardised subscales: (i) Behavioural Regulation Index (BRI) comprising 3 subscores including, Inhibit, Shift, Emotional Control; (ii) Metacognition index (MI) comprising 5 subscores including, Initiate, Working Memory, Plan/Organise, Organisation of Materials, Monitor; as well as a global score called the Global Executive Composite (GEC). | BRIEF GEC<br>BRIEF BRI<br>BRIEF MI |
| | Neuropsychological tests | | | |
| | | Letter-Number Sequencing (WISC-IV, Wechsler (2003)) | | |
| | | | The letter-number sequencing is a working memory task. Sequences of number and letters are read to the participant, and he/she is then asked to re-sequence the numbers in numerical order from lowest to highest and then to sequence the letters in alphabetical order. Standardised scores were used. | Letter-number sequencing |
| | | Tempo Test Rekenen (De Vos, 1992) | | |
| | | | The Tempo Test Rekenen is an arithmetic test consisting of 200 arithmetic number fact problems presented in five rows (one row with addition, one row with subtraction, one row with division, one row with multiplication, and one mixed problem row). Within each row, the problems increase in difficulty. Participant are asked to solve as many items as possible within 1 min per row. The total raw score was age-adjusted for each participant using the procedure described in the main statistical analyses section. | Tempo test |
| | Neurocognitive computerised tasks | | | |
| | | Flanker Visual Filtering Task (Christ, Kester, Bodner, & Miles, 2011) | | |
| | | | The Flanker Visual Filtering Task was used to assess attentional control and information processing speed. Each trial showed a horizontal row of five fish. The participant was asked to respond as quickly as possible to whether the central fish was facing to the left or right. Congruent trials were the ones with all five fish in the horizontal row pointing in the same direction and incongruent trials were the ones with the four distracting fishes pointing in the opposite direction of the central target fish. Reaction time of the congruent condition and | -Flanker processing speed<br>-Flanker inhibition |

| | | |
|---|---|---|
| | of the incongruent condition were used to assess information processing speed, and the inhibition score (reaction time in incongruent conditions – reaction time in congruent conditions) was used as a measure of attentional control. | |
| | Reality Filtering Task (Liverani et al., 2017; Schnider, 2018) | |
| | The Reality Filtering task child-adapted version was used to assess recognition memory and orbitofrontal reality filtering. It consisted of a continuous recognition task composed of two runs with the same picture set but arranged in different order. Accuracy of the second run (D2) and Temporal Context Confusion index (TCC as defined by Schnider, 2018) measures reality filtering. | Reality filtering<br>TCC |
| | **Behaviour and socio-emotional competences** | |
| | Parent questionnaire | |
| | Strength and Difficulties Questionnaire, parent version (SDQ, Goodman (2001)) | |
| | The SDQ parent questionnaire assess overall behaviour problems, emotional symptoms, hyperactivity and inattention, peer relationship problems, and prosocial behaviour. It rates participant's behaviour over the previous 6 months. The SDQ is scored on a Likert scale and includes 25 items, providing a Total Difficulties score. | SDQ total |
| | Self-reported questionnaires | |
| | KIDSCREEN-27 (Robitail et al., 2007) | |
| | The KIDSCREEN-27 is a self-reported questionnaire providing an index of health-related quality of life in children and adolescents. This instrument scored on a Likert scale and includes 27 items, providing a total score. | KIDSCREEN total |
| | Social Goal Scale (SGS, Patrick, Hicks, and Ryan (1997)) | |
| | The SGS is a self-reported questionnaire providing an index of social responsiveness and of goals setting which ultimately gets you involve with some social work. This instrument scored on a Likert scale and includes 11 items providing one score. | Social goal |
| | Self-Compassion Scale – Short form (SCS, Raes, Pommier, Neff, and Van Gucht (2011)) | |
| | The SCS is a self-reported questionnaire comprising 12 items, which produces a total global score that can also be classified into two subscores: negative behaviours toward the self (6 items) or positive behaviours towards the self (6 items). | Self-compassion |
| | Neuropsychological tests | |
| | Affect Recognition (NEPSY-II, Korkman, Kirk, and Kemp (2007) | |
| | The affect recognition subtest assesses the ability to recognise facial emotional expressions (happy, sad, anger, fear, disgust, and neutral) from photographs of children's faces in several matching tasks. In the first task, the participant selected one of the four faces that depicted the same emotion as a child's face at the top of the page. In a second task, the participant selected two photographs of faces that displayed the same affect from a selection of four photographs. Finally, the participant examined a photograph of a child's face for 5 seconds, and then from memory, selected two photographs that matched the same emotion as the face previously shown. Standardised scores were used. | Affect recognition |
| | Theory of Mind (NEPSY-II, Korkman et al. (2007)) | |

| | | | |
|---|---|---|---|
| | | The theory of mind subtest measures understanding of mental functions and other people's perspectives. In the first task, questions are asked to the participant about different verbal scenarios measuring understanding of beliefs, intentions, others' thoughts, ideas and comprehension of figurative language. In the second task, participants have to match facial emotional expressions, from photographs of children's faces, to a scenario. The total raw score was age-adjusted for each participant using the procedure described in the main statistical analyses section. | Theory of mind |

**Supplementary Table S2**. Group comparison on neurobehavioural measures of the VPT and full-term young adolescents.

| | **Preterms Mean (SD)** | **Full-terms Mean (SD)** | **Group comparison** | **Cohen's d** |
|---|---|---|---|---|
| **BRIEF GEC** | **64.28 (9.663)** | **43.36 (17.297)** | **t(36.31)=5.541, p<0.001, q<0.001** | **1.631** |
| **BRIEF MI** | **64.88 (10.826)** | **44.73 (17.398)** | **t(41.03)=5.167, p<0.001, q<0.001** | **1.467** |
| **BRIEF BRI** | **61.91 (14.894)** | **43.23 (18.948)** | **t(42.83)=3.564, p=0.001, q=0.004** | **0.998** |
| Letter-Number sequencing | 10.63 (2.196) | 10.77 (3.054) | t(43.04)=-0.996, p=0.325, q=0.505 | 0.280 |
| Tempo test | 16.19 (5.631) | 17.18 (4.553) | t(48.4)=-1.506, p=0.139, q=0.277 | 0.373 |
| Flanker processing speed | 998.91 (205.635) | 798.61 (366.061) | t(41.51)=1.338, p=0.188, q=0.329 | 0.376 |
| Flanker inhibition | 55.21 (78.926) | 17.36 (62.428) | t(42.93)=1.698, p=0.097, q=0.226 | 0.472 |
| Reality filtering TCC | 0.01 (0.092) | 0.11 (0.298) | t(45.53)=-0.186, p=0.853, q=0.88 | 0.051 |
| **SDQ total** | **12.16 (5.138)** | **6.5 (5.894)** | **t(36.25)=3.186, p=0.003, q=0.01** | **0.938** |
| KIDSCREEN total | 103.13 (22.626) | 90.59 (39.265) | t(32.74)=0.39, p=0.699, q=0.88 | 0.235 |
| Social goal | 3.8 (0.72) | 3.66 (0.952) | t(45.89)=-0.259, p=0.797, q=0.88 | 0.156 |
| **Self-compassion** | **2.74 (0.563)** | **2.91 (0.925)** | **t(34.36)=-2.821, p=0.008, q=0.022** | **0.929** |
| Affect recognition | 10.38 (2.733) | 10.05 (2.87) | t(49.71)=-0.152, p=0.88, q=0.88 | 0.040 |
| Theory of mind | 24.84 (1.706) | 23.55 (5.262) | t(45.51)=0.493, p=0.624, q=0.874 | 0.136 |

Note: Differences between the VPT and full-term groups were examined using independent-sample t test. All p-values that survived false discovery rate (FDR) correction (q < 0.05) are indicated in bold.

**Supplementary Table S3**. Paired-sample t-test on the comparison of neurobehavioural scores before and after MBI intervention in preterm young adolescents.

| | Before MBI mean (sd) | After MBI mean (sd) | Paired-sample t-test | | | | |
|---|---|---|---|---|---|---|---|
| | | | t | df | d | p | q |
| **BRIEF GEC** | **64.28 (9.66)** | **58.2 (10.82)** | **4.673** | **29** | **0.854** | **< 0.001** | **< 0.001** |
| **BRIEF MI** | **64.88 (10.83)** | **56.97 (17.04)** | **5.463** | **29** | **0.452** | **< 0.001** | **< 0.001** |
| BRIEF BRI | 61.91 (14.89) | 58.57 (13.22) | 1.902 | 29 | 0.333 | 0.067 | 0.084 |
| **SDQ total** | **12.16 (5.14)** | **10.5 (4.46)** | **2.379** | **29** | **0.434** | **0.024** | **0.040** |
| Self-compassion | 2.79 (0.47) | 2.95 (0.6) | -1.632 | 28 | -0.303 | 0.114 | 0.114 |

*Note: d shows Cohen's d effect sizes; df degree of liberty; all p-values of paired-sample t-tests that survived false discovery rate (FDR) correction according to Benjamini and Hochberg (1995) are indicated in bold (q < 0.05).*

**Supplementary Table S4.** Mean saliences and their bootstrap-estimated standard deviations for Δ neurobehavioural measures and Δ mean FA of the first PLSC analyses.

| **Salience type: Neurobehavioural functioning** | **Salience (bootstrap estimate standard deviation)** |
|---|---|
| Δ BRIEF GEC | -0.636 (0.073) |
| Δ BRIEF MI | -0.505 (0.117) |
| Δ SDQ Total | -0.025 (0.124) |
| Gestational age | 0.062 (0.125) |
| Age at assessment | -0.457 (0.104) |
| **Salience type: Δ mean FA on extracted white-matter tracts (TrackSeg)** | **Salience (bootstrap estimate standard deviation)** |
| Corpus Callosum - Rostrum | 0.06 (0.062) |
| Corpus Callosum - Rostral body | 0.103 (0.054) |
| Corpus Callosum - Anterior midbody | 0.092 (0.061) |
| Corpus Callosum - Posterior midbody | 0.087 (0.054) |
| Corpus Callosum - Splenium | 0.034 (0.067) |

| | |
|---|---|
| Corticospinal tract_right | 0.092 (0.049) |
| Inferior cerebellar peduncle_left | 0.096 (0.051) |
| Inferior occipito-frontal fascicle_right | 0.044 (0.068) |
| Inferior longitudinal fascicle_left | 0.124 (0.053) |
| Inferior longitudinal fascicle_right | 0.037 (0.067) |
| Optic radiation_left | 0.059 (0.063) |
| Parieto-occipital pontine_left | 0.058 (0.066) |
| Parieto-occipital pontine_right | 0.109 (0.052) |
| Superior longitudinal fascicle I_left | 0.127 (0.045) |
| Superior longitudinal fascicle I_right | 0.089 (0.055) |
| Superior Thalamic Radiation_left | 0.023 (0.056) |
| Uncinate fascicle_right | 0.047 (0.055) |
| Thalamo-parietal_left | 0.083 (0.053) |
| Thalamo-parietal_right | 0.1 (0.049) |
| Striato-fronto-orbital_right | 0.09 (0.048) |
| Arcuate fascicle_left | 0.141 (0.041) |
| Arcuate fascicle_right | 0.129 (0.045) |
| Anterior Thalamic Radiation_left | 0.202 (0.045) |
| Anterior Thalamic Radiation_right | 0.148 (0.043) |
| Corpus Callosum - Genu | 0.164 (0.03) |
| Corpus Callosum - Isthmus | 0.112 (0.049) |
| Cingulum left | 0.138 (0.048) |
| Cingulum _right | 0.137 (0.05) |
| Corticospinal tract_left | 0.14 (0.039) |
| Fronto-pontine tract_left | 0.188 (0.039) |
| Fronto-pontine tract_right | 0.176 (0.046) |
| Inferior cerebellar peduncle_right | 0.123 (0.052) |
| Inferior occipito-frontal fascicle_left | 0.157 (0.052) |
| Middle cerebellar peduncle | 0.156 (0.059) |
| Optic radiation_right | 0.111 (0.051) |
| Superior cerebellar peduncle_left | 0.136 (0.054) |
| Superior cerebellar peduncle_right | 0.177 (0.045) |
| Superior longitudinal fascicle II_left | 0.15 (0.041) |
| Superior longitudinal fascicle II_right | 0.147 (0.035) |

| | |
|---|---|
| Superior longitudinal fascicle III_left | 0.159 (0.044) |
| Superior longitudinal fascicle III_right | 0.122 (0.047) |
| Superior Thalamic Radiation_right | 0.109 (0.048) |
| Uncinate fascicle_left | 0.157 (0.05) |
| Thalamo-premotor_left | 0.153 (0.046) |
| Thalamo-premotor_right | 0.158 (0.05) |
| Thalamo-occipital_left | 0.152 (0.051) |
| Thalamo-occipital_right | 0.12 (0.053) |
| Striato-fronto-orbital_left | 0.169 (0.043) |
| Striato-premotor_left | 0.167 (0.036) |
| Striato-premotor_right | 0.118 (0.049) |

**Supplementary Table S5.** Mean saliences and their bootstrap-estimated standard deviations for Δ neurobehavioural measures and Δ mean ICVF and ODI of the second PLSC analyses.

| **Salience type:<br>Neurobehavioural functioning** | **Salience (bootstrap estimate standard deviation)** |
|---|---|
| Δ BRIEF GEC | -0.689 (0.068) |
| Δ BRIEF MI | -0.603 (0.103) |
| Δ SDQ Total | -0.032 (0.11) |
| Gestational age | -0.04 (0.08) |
| Age at assessment | -0.262 (0.099) |
| **Salience type:<br>Δ mean ICVF on extracted white-matter tracts (TrackSeg)** | **Salience (bootstrap estimate standard deviation)** |
| Arcuate fascicle left | -0.076 (0.032) |
| Arcuate fascicle right | 0.009 (0.038) |
| Anterior Thalamic Radiation left | -0.098 (0.057) |
| Anterior Thalamic Radiation right | -0.047 (0.04) |
| Corpus callusum - Rostrum | -0.029 (0.043) |

| | |
|---|---|
| Corpus callusum -Genu | -0.052 (0.041) |
| Corpus callusum -Rostral body | -0.014 (0.048) |
| Corpus callusum -Anterior midbody | 0.009 (0.04) |
| Corpus callusum -Posterior midbody | -0.011 (0.055) |
| Corpus callusum -Isthmus | -0.031 (0.044) |
| Corpus callusum -Splenium | -0.035 (0.05) |
| Cingulum left | -0.046 (0.052) |
| Cingulum right | -0.023 (0.039) |
| Corticospinal tract left | -0.039 (0.044) |
| Corticospinal tract right | 0.007 (0.034) |
| Fronto-pontine tract left | -0.035 (0.037) |
| Fronto-pontine tract right | -0.003 (0.035) |
| Inferior cerebellar peduncle left | -0.101 (0.046) |
| Inferior cerebellar peduncle right | -0.014 (0.045) |
| Inferior occipito-frontal fascicle left | -0.054 (0.045) |
| Inferior occipito-frontal fascicle right | 0.064 (0.049) |
| Inferior longitudinal fascicle left | -0.01 (0.049) |
| Inferior longitudinal fascicle right | 0.061 (0.045) |
| Middle cerebellar peduncle | -0.023 (0.037) |
| Optic radiation left | -0.086 (0.053) |
| Optic radiation right | 0.005 (0.048) |
| Parieto-occipital pontine left | -0.006 (0.044) |
| Parieto-occipital pontine right | 0.036 (0.049) |
| Superior cerebellar peduncle left | -0.018 (0.038) |
| Superior cerebellar peduncle right | 0.031 (0.045) |
| Superior longitudinal fascicle I left | -0.063 (0.041) |
| Superior longitudinal fascicle I right | -0.03 (0.04) |
| Superior longitudinal fascicle II left | -0.066 (0.036) |
| Superior longitudinal fascicle II right | 0.018 (0.035) |
| Superior longitudinal fascicle III left | -0.026 (0.044) |
| Superior longitudinal fascicle III right | 0.041 (0.045) |
| Superior Thalamic Radiation left | -0.072 (0.046) |
| Superior Thalamic Radiation right | -0.014 (0.034) |
| Uncinate fascicle left | -0.052 (0.049) |

| | |
|---|---|
| Uncinate fascicle right | -0.018 (0.036) |
| Thalamo-premotor left | -0.039 (0.052) |
| Thalamo-premotor right | -0.013 (0.057) |
| Thalamo-parietal left | -0.12 (0.047) |
| Thalamo-parietal right | 0.014 (0.051) |
| Thalamo-occipital left | -0.103 (0.036) |
| Thalamo-occipital right | 0.007 (0.043) |
| Striato-fronto-orbital left | -0.028 (0.044) |
| Striato-fronto-orbital right | 0.001 (0.035) |
| Striato-premotor left | -0.08 (0.045) |
| Striato-premotor right | -0.032 (0.053) |
| **Salience type: Δ mean ODI on extracted white-matter tracts (TrackSeg)** | **Salience (bootstrap estimate standard deviation)** |
| Arcuate fascicle left | -0.142 (0.029) |
| Arcuate fascicle right | -0.108 (0.037) |
| Anterior Thalamic Radiation left | -0.157 (0.025) |
| Anterior Thalamic Radiation right | -0.136 (0.031) |
| Corpus callusum - Rostrum | -0.099 (0.034) |
| Corpus callusum -Genu | -0.137 (0.023) |
| Corpus callusum -Rostral body | -0.08 (0.041) |
| Corpus callusum -Anterior midbody | -0.051 (0.054) |
| Corpus callusum -Posterior midbody | -0.067 (0.044) |
| Corpus callusum -Isthmus | -0.093 (0.054) |
| Corpus callusum -Splenium | -0.027 (0.059) |
| Cingulum left | -0.119 (0.037) |
| Cingulum right | -0.106 (0.032) |
| Corticospinal tract left | -0.155 (0.03) |
| Corticospinal tract right | -0.07 (0.04) |
| Fronto-pontine tract left | -0.186 (0.022) |
| Fronto-pontine tract right | -0.145 (0.033) |
| Inferior cerebellar peduncle left | -0.123 (0.049) |
| Inferior cerebellar peduncle right | -0.135 (0.052) |

| | |
|---|---|
| Inferior occipito-frontal fascicle left | -0.125 (0.05) |
| Inferior occipito-frontal fascicle right | -0.067 (0.064) |
| Inferior longitudinal fascicle left | -0.107 (0.046) |
| Inferior longitudinal fascicle right | -0.012 (0.054) |
| Middle cerebellar peduncle | -0.166 (0.047) |
| Optic radiation left | -0.093 (0.052) |
| Optic radiation right | -0.093 (0.055) |
| Parieto-occipital pontine left | -0.031 (0.05) |
| Parieto-occipital pontine right | -0.077 (0.06) |
| Superior cerebellar peduncle left | -0.117 (0.044) |
| Superior cerebellar peduncle right | -0.182 (0.038) |
| Superior longitudinal fascicle I left | -0.104 (0.036) |
| Superior longitudinal fascicle I right | -0.077 (0.041) |
| Superior longitudinal fascicle II left | -0.129 (0.031) |
| Superior longitudinal fascicle II right | -0.121 (0.029) |
| Superior longitudinal fascicle III left | -0.148 (0.027) |
| Superior longitudinal fascicle III right | -0.083 (0.044) |
| Superior Thalamic Radiation left | -0.138 (0.03) |
| Superior Thalamic Radiation right | -0.11 (0.041) |
| Uncinate fascicle left | -0.137 (0.028) |
| Uncinate fascicle right | -0.102 (0.034) |
| Thalamo-premotor left | -0.135 (0.027) |
| Thalamo-premotor right | -0.102 (0.032) |
| Thalamo-parietal left | -0.132 (0.043) |
| Thalamo-parietal right | -0.105 (0.054) |
| Thalamo-occipital left | -0.11 (0.039) |
| Thalamo-occipital right | -0.086 (0.051) |
| Striato-fronto-orbital left | -0.033 (0.04) |
| Striato-fronto-orbital right | -0.078 (0.051) |
| Striato-premotor left | -0.152 (0.039) |
| Striato-premotor right | -0.072 (0.049) |

### Supplementary Figures

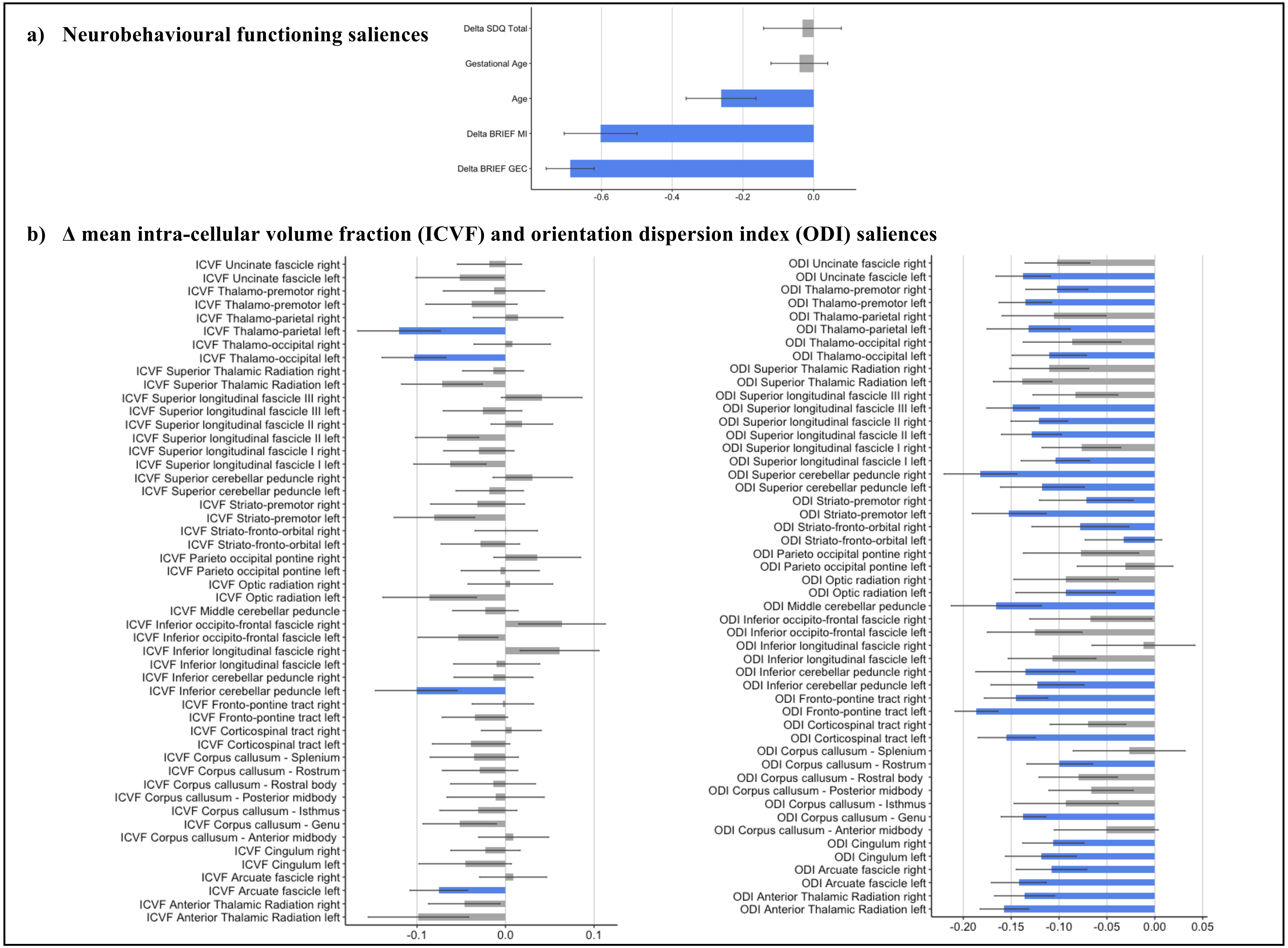

**Figure S1.** Illustration of the results from the second PLSC analyses. a) Neurobehavioural functioning saliences: the diverging graph show mean saliences averaged across bootstrap samples and their bootstrap-estimated standard deviations (x-axis) for each neurobehavioural functioning measure (y-axis); robust saliences are represented in blue; of note, saliences bellow 0 indicate a decreased in scores after MBI and saliences above 0 indicate an increased in scores after MBI. b) $\Delta$ mean intra-cellular volume fraction (ICVF) and orientation dispersion index (ODI) saliences: the diverging graph show mean saliences averaged across bootstrap samples and their bootstrap-estimated standard deviations (x-axis) for each $\Delta$ mean ICVF and ODI along a given tract (y-axis); the tracts extracted from TractSeg and showing robust $\Delta$ mean ICVF or ODI saliences are represented in blue; of note, saliences below 0 indicate a decreased in scores after MBI and saliences above 0 indicate an increased in scores after MBI.
